# Biological aging measured from electronic health records is associated with recovery following cardiothoracic surgery

**DOI:** 10.64898/2026.10.16.26365756

**Authors:** CP Ryan, J Jung, P Kurlansky, W Marella, H Wang, VK Moitra, DW Belsky

## Abstract

Biological aging, the decline in physiological resilience that increases our vulnerability to disease and death as we grow older, can be measured from standard clinical-laboratory panels. Measurement of biological aging from laboratory data embedded in electronic health records (EHRs) could enhance precision healthcare. However, suitability of EHR-embedded laboratory records for this purpose is uncertain. We evaluated feasibility of EHR biological age measurement in a cohort of n=4,313 cardiothoracic-surgery patients at a large US Hospital and tested whether measured biological aging was associated with patient outcomes. Patients who were biologically older prior to surgery more often died or experienced adverse outcomes as compared to those measured to be biologically younger. Postoperative biological-aging trajectories were associated with length of intensive-care-unit (ICU) stays and in-hospital death. EHR laboratory data provide an opportunity for scalable implementation of biological aging measurement within healthcare systems, with potential to enhance patient monitoring and risk stratification.

## Main

Patients entering clinical care differ in their physiological function and vulnerability to acute stress. This heterogeneity has direct consequences for clinical risk: patients facing the same illness or undergoing the same procedure can experience markedly different outcomes. Chronological age is a well-established risk factor underlying heterogeneity in vulnerability,^1^ but it is incomplete. Individuals of the same age differ in their underlying biological condition and risks for adverse outcomes. Geroscience provides a framework for understanding this variation by characterizing aging as a biological process that progresses at different rates in different individuals.^2,3^ There are now methods to quantify individual differences in biological aging, including methods that rely on data from standard clinical laboratory panels.^4,5^ These methods could be applied to implement biological aging measurement within electronic health records (EHRs), opening a path toward increased precision in patient risk stratification as well as offering opportunities for population surveillance and health services research.

In studies across the US, Europe, and Asia, individuals whose laboratory values show they are biologically old relative to their chronological age are at increased risk of incident chronic disease and disability and have shorter lifespans as compared with age-peers who look biologically younger.^6–10^ Even in younger people who are mostly free of chronic disease, differences in measured biological aging correspond to early-emerging deficits in physical and cognitive functions^11^. Across the lifespan, biological age measurements correlate with a range of healthy lifestyle factors as well as social determinants of health.^12–15^ Finally, these measurements are responsive to calorie restriction – a gold standard intervention to slow aging in animals – in healthy, non-obese adult humans.^16,17^ Despite this progress, and despite ready availability of data within EHRs, biological aging measurement in healthcare remains limited.

Two key knowledge gaps limit translation of biological aging metrics from research studies into tools for precision healthcare. First, biological-age methods for clinical laboratory test data were developed in community-based samples;^18^ it is unknown if values are interpretable or informative in clinical settings characterized by acute illness, physiological stress, and medical intervention. Second, biological-age methods for clinical laboratory test data were developed in research cohorts; it is unknown if biological age can be reliably quantified from laboratory tests completed as part of clinical care.^19,20^ Addressing these gaps is critical for determining whether biological aging metrics can be translated from population studies into real-world clinical contexts.

To address these gaps, we conducted a study of an established approach to measure biological aging from clinical laboratory data using EHR data from a large academic health system. We analyzed data from cardiothoracic surgery patients at Columbia University Irving Medical Center (CUIMC). We tested how biological aging measured from perioperative EHR data could inform prediction of short-term postoperative outcomes. We conducted two sets of analyses. First, we analyzed laboratory data collected during the 6 months prior to surgery and focused on prediction of surgical outcomes and 30-day post-operative survival. Second, we analyzed laboratory tests collected in the 48 hours immediately following surgery to test associations with length of stay in the intensive care unit (ICU) and in-hospital mortality. Together, these analyses provide a practical test of whether biological age can be measured within EHR systems and used to characterize risk and recovery in a real-world clinical setting.

## Results

### Pre-operative biological age and surgical outcomes

We analyzed linked Society of Thoracic Surgeons Adult Cardiac Surgery Database (STS ACSD) and electronic health record (EHR) data for patients who underwent cardiothoracic surgery at New York-Presbyterian Hospital/Columbia University Irving Medical Center (NYP/CUIMC) between February 3, 2020, and November 16, 2024. The linked study extract included 4,313 cases. We measured preoperative biological age using laboratory results accumulated during the six months preceding the index surgery for 3,343 patients across 11,625 measurement occasions (**Fig. 1**). Briefly, we identified laboratory panels available for most patients (a basic metabolic panel, a hepatic function panel, and a complete blood count panel with platelets and no differential measure for leukocyte subtypes) and used the BioAge software^17^ and data from NHANES III (1988-1994) to parameterize a biological age algorithm from these panels using the PhenoAge method **(Methods and Supplementary Methods; Tables S1-S7; Figures S1-S6**).^18^ A descriptive summary of the patients in the New York Presbyterian Hospital cardiothoracic surgery record system with preoperative laboratory records sufficient to calculate the biological aging metric are shown in **Table S8**. Patients’ biological age values were highly correlated with their ages at laboratory testing (r=0.82), although values had a negative bias; patients biological ages were on average 3.3 years younger than their calendar ages (**Fig. 2**). This negative bias in biological ages suggests that our patient cohort measured during the 2020s was aging more slowly as compared with the US population in the late 1980s and early 1990s. But we cannot rule out changes in laboratory methods affecting distributions of assay values as a cause.

**Figure 1.**
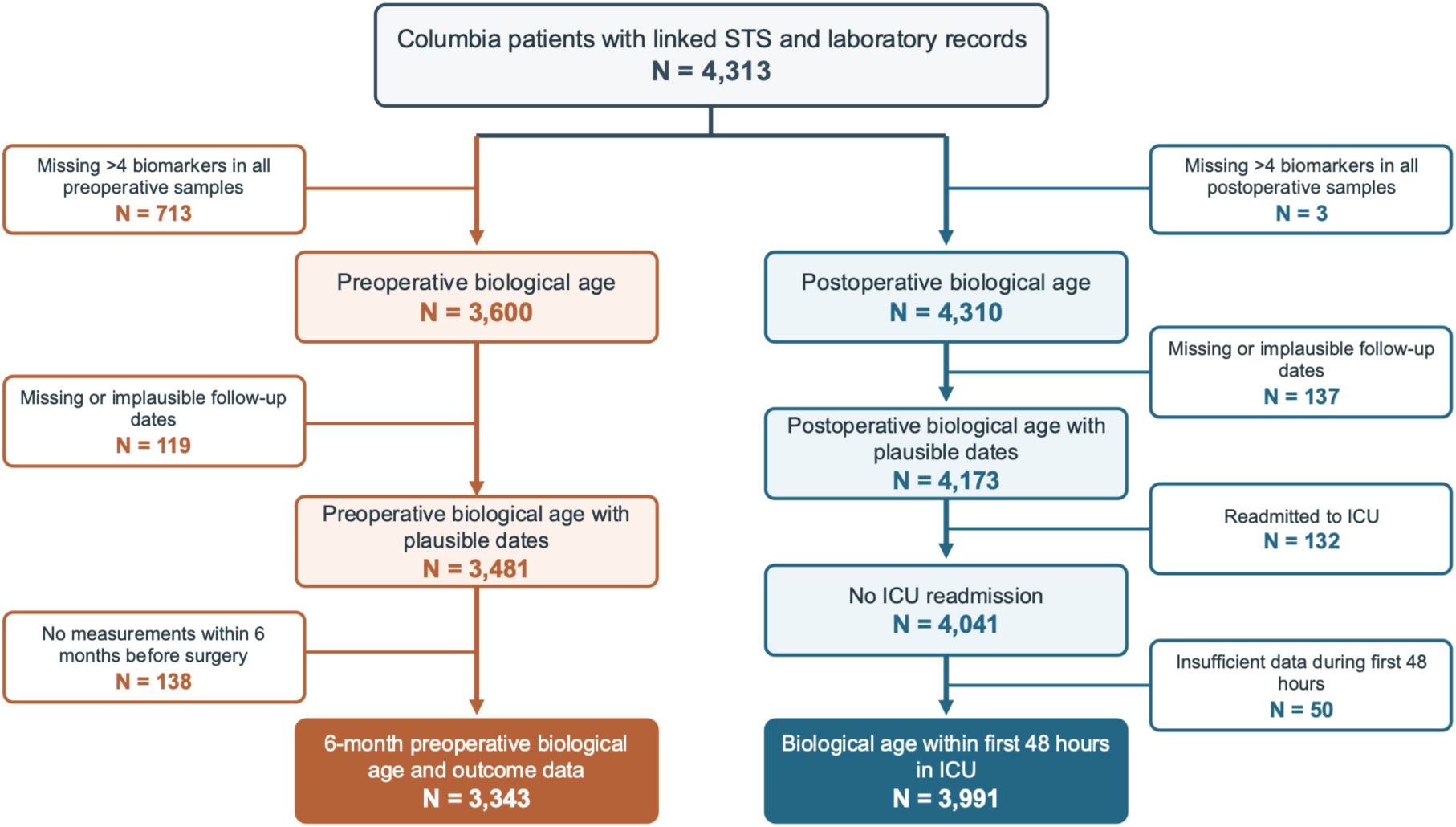
**Cohort flow for pre-operative and post-operative biological age analyses using electronic records from New York Presbyterian Hospital.** Complete demographic, operative, and laboratory record information was available for n=4,313 patients. Patients were then stratified into pre-operative and post-operative biological age analyses. For the pre-operative analysis, exclusions were applied for missing biomarker data, implausible or missing follow-up dates, and absence of laboratory measurements within six months prior to surgery, yielding a final analytic sample of 3,343 patients with both pre-operative biological age and operative outcome data. For the post-operative analysis, patients were excluded for missing biomarker data, implausible follow-up dates, ICU readmission, missing laboratory data within the first 48 hours, or missing discharge information, resulting in 3,991 patients with post-operative 48-hour ICU biological age and discharge mortality status.

**Figure 2.**
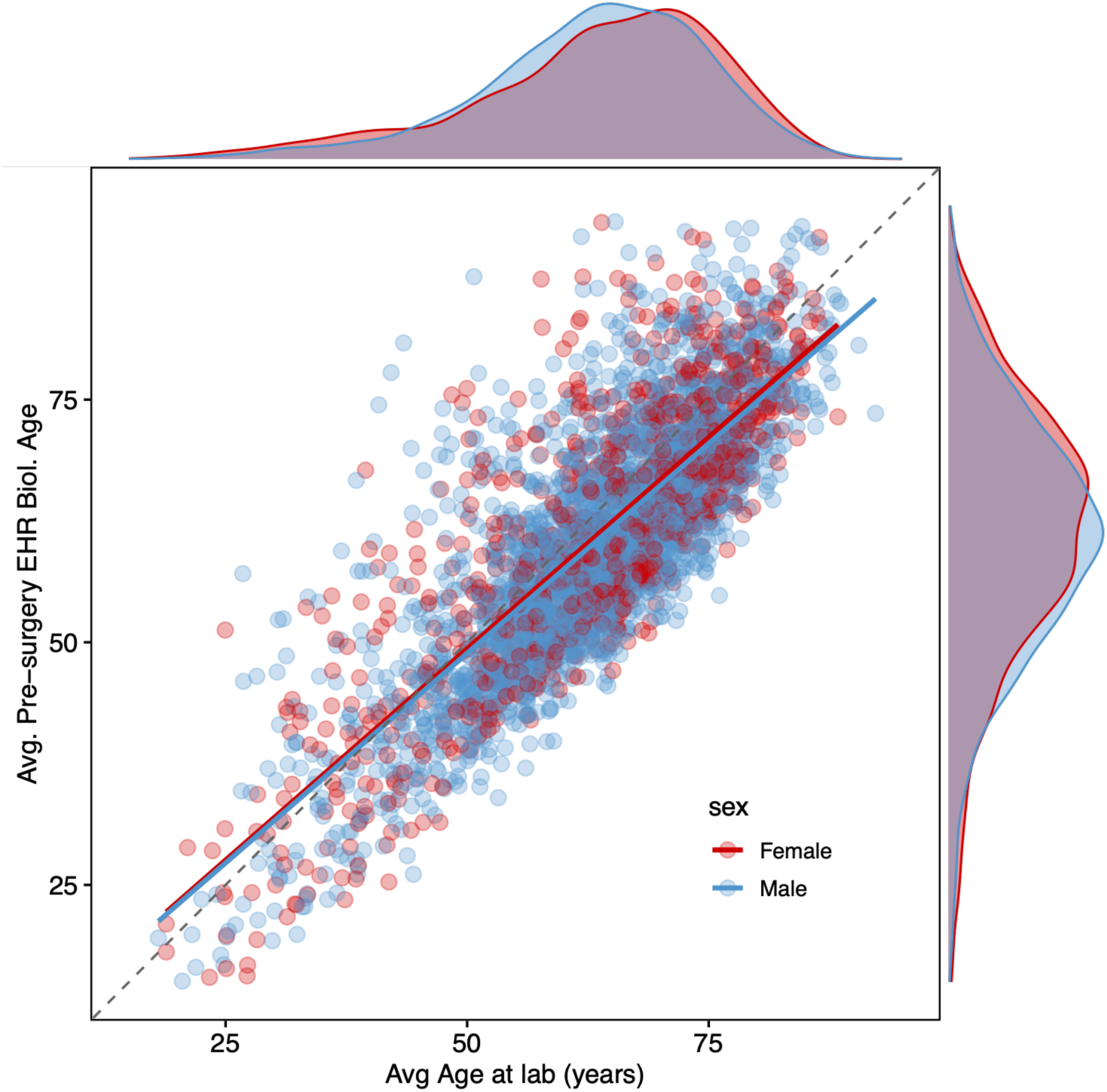
**Patients’ biological age plotted against their chronological age at laboratory testing.** The figure shows a scatterplot of average 6-month pre-operative biological age versus average 6-month chronological age at lab measurement, colored by sex (males, blue; females, red), with each point representing an individual patient (n=3,343). The dashed, gray line represents the identity function, where biological age and chronological age are equal. Solid, colored lines represent a simple regression between the two axes colored by sex. Density plots show distributions of average at lab (x-axis) and average 6-month pre-operative biological age (y-axis). Patients’ biological age values were highly correlated with their ages at laboratory testing, although they tended to be slightly younger biologically than their chronological age.

For analysis, we calculated the difference between a patient’s biological age and their calendar age (BA-CA). We then scaled this value based on the population distribution NHANES III data used to develop the metric. A one-unit increment in biological aging in our analysis corresponds to a one standard deviation difference in the NHANES.

*Patients with older pre-operative biological ages were at increased risk for adverse outcomes* We tested if patients who were biologically-older prior to surgery were likely to experience negative outcomes (30-day mortality [n=3,343]; stroke, renal failure, prolonged ventilation, sepsis, or life-threatening complications [ns=1,617-3,342]; **Methods and Supplementary Methods, Table S1**) compared to patients who were biologically-younger. Patients with older pre-operative biological ages were more likely to die over the 30 days following surgery compared to those with younger biological ages (risk ratio per standard deviation [RR]=1.61, 95% CI: 1.46-1.78, p<0.001). They also experienced adverse outcomes and complications more often than those with younger biological ages (RR range 1.12-1.54; p<0.001 for all; **Table S9**). Effect-sizes were similar for men and women (**Figure 3** and **Table S9)**.

**Figure 3.**
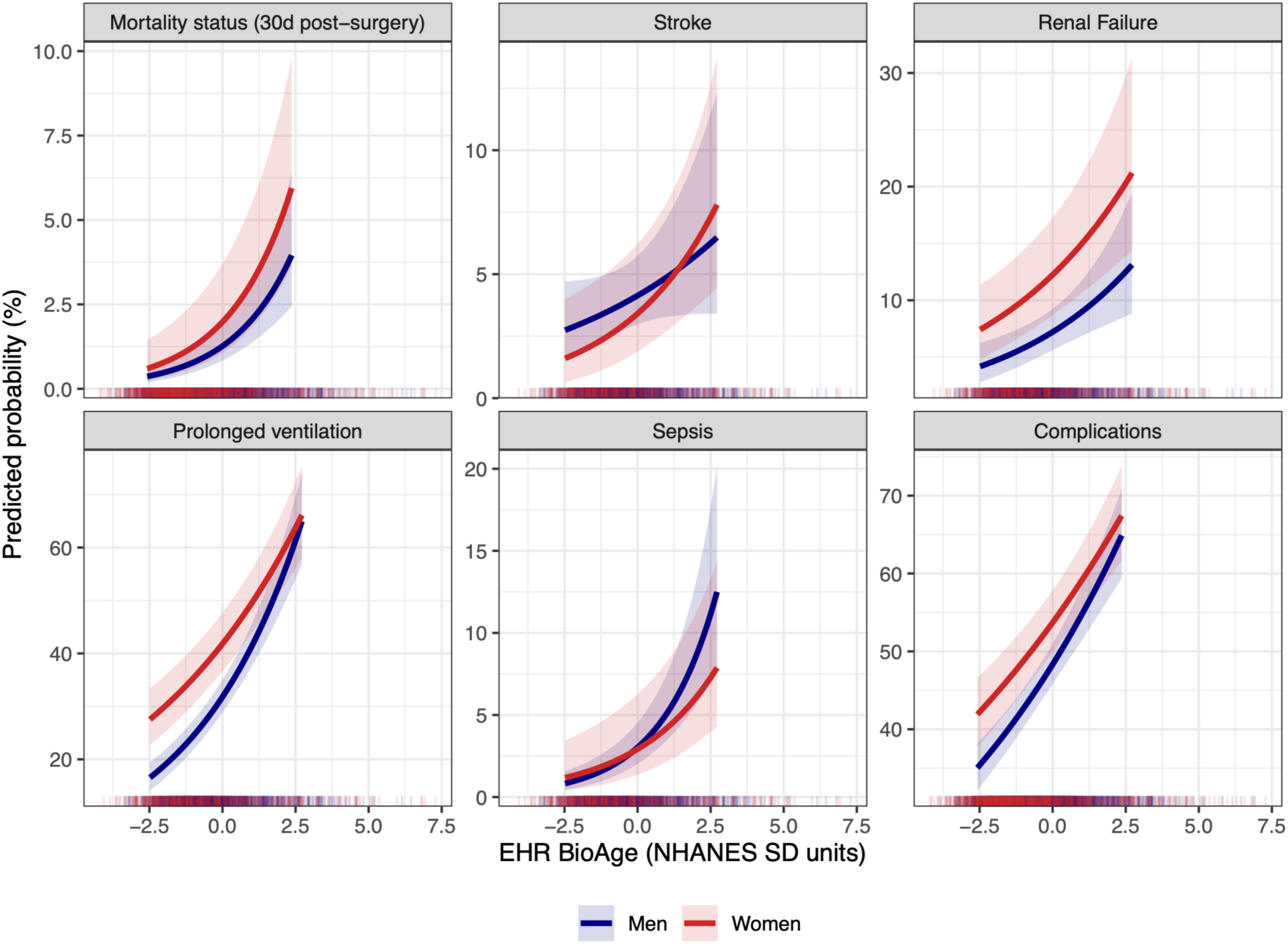
**Predicted postoperative outcome probability across EHR BioAge for men and women.** Predicted probabilities of postoperative outcomes are shown across the 5th–95th percentile range of preoperative EHR BioAge, expressed in NHANES standard deviation units. Predicted probabilities were estimated from sex-stratified Poisson regression models with robust standard errors, adjusted for chronological age and age-squared. Lines show predicted probabilities for men and women separately; shaded bands indicate 95% confidence intervals. Rug marks along the x-axis show the distribution of observed EHR BioAge values in the analytic sample and may extend beyond the fitted prediction lines because the prediction curves are restricted to the central 5th–95th percentile range. Outcomes shown are 30-day post-surgery mortality (n_total_=3343, n_events_=61), stroke (n_total_=1617, n_events_=61), renal failure (n_total_=1617, n_events_=129), prolonged ventilation (n_total_=1617, n_events_=557), sepsis (n_total_=1617, n_events_=59), and complications of surgery (n_total_=3342, n_events_=1617).

### EHR Biological Age captures similar information as established clinical risk scores

An established approach to evaluating a patient’s risk of adverse outcomes of surgery come from The Society of Thoracic Surgeons (STS) risk prediction models^22,23^. These models score a patient from 0-1 based on probability of experiencing a range of negative outcomes (e.g. stroke, renal failure, prolonged ventilation, sepsis, morbidity, and life-threatening complications) based on detailed pre-operative information including patient demographics, health behaviors, surgical procedure, prevalent comorbidities, and laboratory chemistries at the time of surgery^22,23^ (**Figure S7**). The STS risk score is manually curated and only calculated in a subset of patients. In the database we analyzed, STS risk scores were available for about two thirds of patients (60-70% depending on outcome) of patients (comparison of patients with and without STS risk scores in **Table S10**).

We tested if biological age information captured a similar dimension of patient risk to these established risk scores. Patients’ pre-operative EHR Biological Age values were correlated with but distinct from their STS risk scores (r=0.26-0.46; **Figure S8**). In the subset of patients with both biological aging and STS risk score data, we found that combining information from both metrics improved risk classification only in the case of sepsis (**Figure S9; Tables S11-12**).

### Post-operative biological age and intensive-care-unit outcomes

We next asked whether biological aging measured during the 48h following surgery could help identify patients at risk for adverse outcomes (**Table S13**). A descriptive summary of the patients in the New York Presbyterian Hospital cardiothoracic surgery record system with postoperative laboratory records sufficient to calculate the biological aging metric are shown in **Table S8**. In the ICU, patients’ lab values are calculated regularly. On average, patients in our sample contributed on 3.0 (SD=1.2, range=1-10) measurements over the 48h post-operative period. Trajectories were variable, but exhibited a stereotypical pattern of reduction relative to pre­operative biological age followed by recovery back toward pre-operative age (**Fig. S10**), driven by changes in blood analytes known to vary with acute physiological stress (e.g. glucose and bilirubin increased following surgery and then attenuated toward pre-operative values; **Fig. S11-S13**).

We quantified differences in patients’ trajectories in two ways: (i) we averaged biological aging measurements over the first 48 hours after surgery, and (ii) we computed slopes of change in biological aging over the same time period.

#### Average post-operative biological age is associated with ICU length of stay, chronic critical illness, and in-hospital mortality

Patients measured to be biologically older in the 48h following surgery tended to have longer ICU stays (n=3,991 patients with 12,091 biological age measurements; discharge HR=0.64 per biological aging SD, 95% CI 0.62-0.67), more often experienced stays >7d, classified as “chronic critical illness” (odds ratio [OR]=1.13, 95% CI=1.11-1.14, p<0.001), and were more likely to die prior to discharge (OR=1.16, 95% CI=1.13-1.19, p<0.001). Details of these analyses are reported in **Figure 4** and **Table S14**.

**Figure 4.**
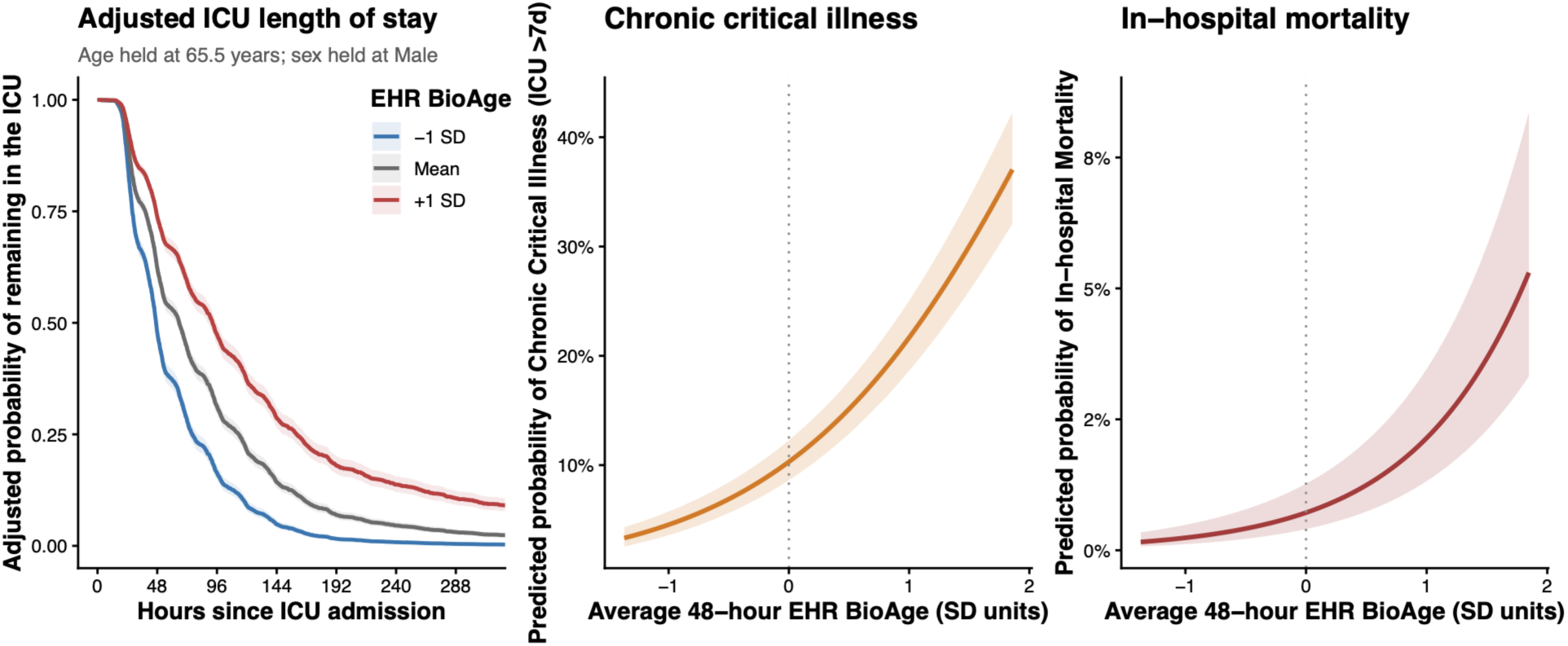
**Postoperative outcomes across average 48-hour post-op EHR BioAge.** (left) Cox model–estimated probability of remaining in the ICU at average EHR BioAge values of −1 SD, the mean, and +1 SD. ICU discharge was modeled as the event; patients who died in the ICU were right-censored at the time of death. Adjusted predicted probabilities of prolonged ICU stay (>7 days, middle) and in-hospital mortality (right) across continuous average 48-hour EHR BioAge. EHR BioAge was calculated from measurements collected during the first 48 hours of ICU care. All models were adjusted for age at surgery, age squared, and sex. Shaded bands indicate 95% confidence intervals, and continuous EHR BioAge values are expressed in standard deviation units.

#### Post-operative biological age trajectories are associated with ICU length of stay, chronic critical illness, and in-hospital mortality

We analyzed trajectories of patient biological age values over the 48h following surgery using mixed models with random linear and quadratic slope terms. We then tested how patients’ baseline biological ages and their linear and quadratic slopes of change related to their lengths of stay and risk of death.

Consistent with analysis of 48h average biological age, patients with older baseline biological age were at increased risk for longer stays and in-hospital mortality (discharge HR=0.69, 95% CI=0.67–0.71, p<0.001; prolonged ICU stay OR=2.24, 95% CI=2.05-2.46, p<0.001; death before discharge OR=2.87, 95% CI=2.34-3.54, p<0.001) (**Fig. S14**). Patients’ slopes of change were also related to adverse outcomes. Specifically, accelerating increases in biological aging during the 48h following surgery were associated with longer stays and increased risk of death (**Fig. S14**; **Table S15**).

#### Biological aging dynamics in the 48h following surgery improved risk classification for chronic critical illness and in-hospital mortality

We compared risk classification for chronic critical illness and in-hospital mortality based on demographic information (age and sex) to classification that included information about biological aging using ROC analysis. Demographics-only models achieved AUCs of 0.56 for chronic critical illness and 0.66 for in-hospital mortality. Incorporation of biological aging trajectory parameters improved discrimination to AUC=0.75 for chronic critical illness (95% CI 0.73-0.77) and to AUC=0.85 for in-hospital mortality (95% CI 0.81-0.90). Full results are reported in **Figure S15** and **Table S16**.

## Discussion

We analyzed electronic health records (EHR) from a cohort of cardiothoracic surgery patients at a large teaching hospital in New York City to test the translational potential of laboratory­chemistry biological aging measurements for precision healthcare. In research datasets, these measurements are predictive of chronic disease incidence, disability, and mortality. But their potential to support healthcare decision making is uncertain. We used established methods and training data^24^ to develop a biological age measurement customized to available health-system data. We then used patients’ EHR data to calculate their pre-operative biological age and to track changes in their biological ages over the 48h period following surgery. We found that patient biological ages calculated from pre-operative EHR data helped distinguish risk for short­term adverse surgical outcomes including stroke, renal failure, sepsis, life-threatening or long­term complications, and mortality among patients who were the same sex and calendar age. Further, patient post-operative biological ages calculated from intensive-care-unit (ICU) data embedded in the EHR were associated with length of stay and in-hospital mortality.

These findings contribute evidence that geroscience-informed measurements of biological aging have translational potential in healthcare settings. Critically, they affirm the suitability of passively collected laboratory records embedded in EHRs for calculation of metrics developed in community-based research cohorts^25^. The pre-operative laboratory values in the database we analyzed were generated for a range of purposes, including to monitor prevalent health conditions. Even so, when we imposed a model developed in a community-based research cohort, the results were interpretable and associated with important patient outcomes. The same was true of biological ages calculated from laboratory data accumulated during the immediate post-operative period, when patients were recovering in the ICU.

Records of laboratory test results are now accumulated at scale and curated within harmonized databases at the health-system level.^25,26^ EHR-based surveillance of biological aging has the potential to integrate geroscience into routine clinical care and precision medicine.^19^ In routine clinical care, the simplicity of a biological age may help providers communicate with patients about health trajectories, including in response to changes in behavior, lifestyle, or clinical management of a chronic condition. Within the scope of precision medicine, biological ages can help care providers tailor screening and treatment to a patient with greater specificity than is afforded by their demographic characteristics.^20,26^

There are limits to what a simple biological aging measurement can accomplish. In the subset of patients in our cohort for whom Society for Thoracic Surgery risk scores^22,23^ were available (roughly two-thirds), biological aging measurements offered little additional information.

However, these risk scores require manual curation of clinical, surgical, physiological, and lifestyle features selected to predict specific outcomes (e.g. sepsis, stroke) and are not available for many patients. They are procedure-specific risk predictions, not general-purpose patient measurements. Moreover, because the risk scores are based on proprietary algorithms, we were unable to determine if they could be enhanced by incorporation of biological aging measurements. If and when such measurements become part of EHRs, it will be feasible to consider their integration into existing risk stratification tools.

We acknowledge limitations. Our study was based on data from a cohort of surgical patients in a single health system. Replication in other health systems is needed to establish robustness of the result. Similarly, parallel analysis of other patient cohorts and outcomes is needed to confirm that the result generalizes beyond the context of cardiothoracic surgery. Within the Columbia surgical cohort, biological age analyses required sufficient biomarker measurements during the relevant perioperative period. The preoperative analysis was further limited to patients with measurements within six months before surgery. While small in number, patients included in these analyses may therefore differ from those without sufficient measurements, particularly if preoperative testing occurred outside the health system. Our biological age measure was also constrained by the laboratory panels available in the EHR. Certain parameters that have proven useful in research cohorts, such as red-cell-distribution width (RDW), were less widely available. Our results therefore represent a conservative estimate of the value of geroscience-informed measurements.

Within the context of these limitations, our study establishes proof of concept for integration of geroscience-based measurements of biological aging into EHRs. Our findings show that biological aging measurement is feasible with data passively accumulated in patients’ EHRs and that these measurements have potential to inform precision medicine approaches to patient care. Age is among the most powerful risk factors in medicine. But it is known to be imprecise. Incorporating biological aging measurements into patient EHRs can furnish healthcare providers with important knowledge about patient heterogeneity without requiring additional testing.

## Methods

### Study population and data sources

#### New York Presbyterian (NYP) Hospital electronic health record database

We conducted a retrospective cohort study using electronic health record (EHR) data from patients admitted to the cardiothoracic intensive care unit at New York-Presbyterian Hospital/Columbia University Irving Medical Center (NYP/CUIMC) between February 3^rd^, 2020 and November 16^th^, 2024. The study population included all adult patients who underwent cardiothoracic surgery during the study period and were captured in the Society of Thoracic Surgeons Adult Cardiac Surgery Database (STS ACSD) at our institution.

Clinical and perioperative variables were obtained from the institutional STS database, which contains prospectively collected and manually adjudicated data. These records were linked to longitudinal EHR data extracted from the Epic Clarity database providing structured clinical information including laboratory measurements, vital signs, and encounter-level data for research purposes. Data linkage was performed using record identifiers unique each patient.

The study was approved by the Columbia University Irving Medical Center Institutional Review Board, with a waiver of informed consent due to the retrospective nature of the analysis and minimal risk to participants.

NYP demographic, operative, and discharge data.

Complete demographic, operative, and laboratory records were available for n=4,313 patients (**Fig. 1**). Preoperative laboratory data sufficient to calculate biological age were available for n=3,343 patients (**Fig. 1**). Short-term postoperative outcomes included stroke, renal failure, prolonged ventilation, sepsis, and complications that could cause life-threatening or long-term deficits during hospitalization. Intensive care unit stay was measured in hours from exit from the operating room. Mortality records documented in-hospital death and mortality status 30 days after surgery (**Table S1**).

Biological age construction and projection

We defined our biological age algorithm in 3 steps. In the first step, we ingested laboratory records embedded in EHRs for all patients with an index surgery between 2020-02-03 and 2024-11-16. We reviewed records for all laboratory tests available in the database as of 2025­02-07, and identified metabolic, liver, and hematology panels that common enough to be measured at least one time in all patients. The EHR laboratory data and our approach to panel selection and data preparation are described the **Supplementary Methods**.

In the second step, developed a custom biological age algorithm based on the laboratory tests available in the EHR database. We analyzed data from the National Health and Nutrition Examination Survey (NHANES) III (1988–1994). We followed the approach introduced with PhenoAge^18^. This approach involves two steps. First, a hazard model is used to integrate biomarker information into a mortality risk prediction model. Second, a separate hazard model is fitted to the age distribution to create a crosswalk from mortality risk to the age at which that risk is typical for the population. Together, the risk prediction model and the crosswalk form the biological age algorithm. The first piece estimates mortality risk. The second piece converts that risk into a biological age value. Within a population, people with older biological age relative to their chronological age are at greater risk for all-cause mortality, whereas people with younger biological age than their chronological age are at a lower risk of mortality.

In our analysis, we fit an elastic-net Cox regression^28^ to the biomarker panels and survival data accumulated over on average 21.9 years of follow-up (SD=8.6y, range=0.1-31.1y; N=12,283, deaths=4,968). This analysis selected 18 laboratory tests (alanine aminotransferase, albumin, aspartate aminotransferase, alkaline phosphatase, bilirubin, blood urea nitrogen, calcium, chloride, creatinine, globulin, glucose, mean cell hemoglobin, mean cell volume, mean platelet volume, potassium level, red blood cell count, red cell distribution width, and white blood cell count) in addition to chronological age (**Table S4**). We then re-fit models separately in women and men to define sex-specific biological age algorithms (**Table S5**). These algorithms delivered comparable prediction of mortality to the original PhenoAge in independent data from NHANES IV (N=47,524, deaths=6,376). Detailed methods and results from biological age algorithm construction and validation in NHANES are described in the **Supplementary Methods**.

In the third step, we used the BioAge package^24^ to project our new algorithm onto the EHR laboratory test data using the 18 analytes and coefficients selected in NHANES. Our imputation step, which involved adding the NHANES mean for a given biomarker if an individual was missing 4 or fewer of the 18 analytes, resulted in a total of 18,586 instances of biological age across n=3600 patients prior to surgery, and 62,504 instances of biological age across n=4310 patients after surgery (**Fig. 1**). A comparison all patients in the database (n=17,635) with the subset of this cohort with pre-op laboratory panels is shown in **Table S7**. Additional details on EHR BioAge construction and analysis are described in the **Supplementary Methods**.

## Data Availability

The study uses restricted patient-level data that cannot be shared or transferred outside the approved institutional environment.

## Acknowledgement

This research received support from the Columbia University Irving Medical Center Healthy Aging Initiative (CHAI) Pilot Grant.

## Supplementary Tables

**Table S1.**
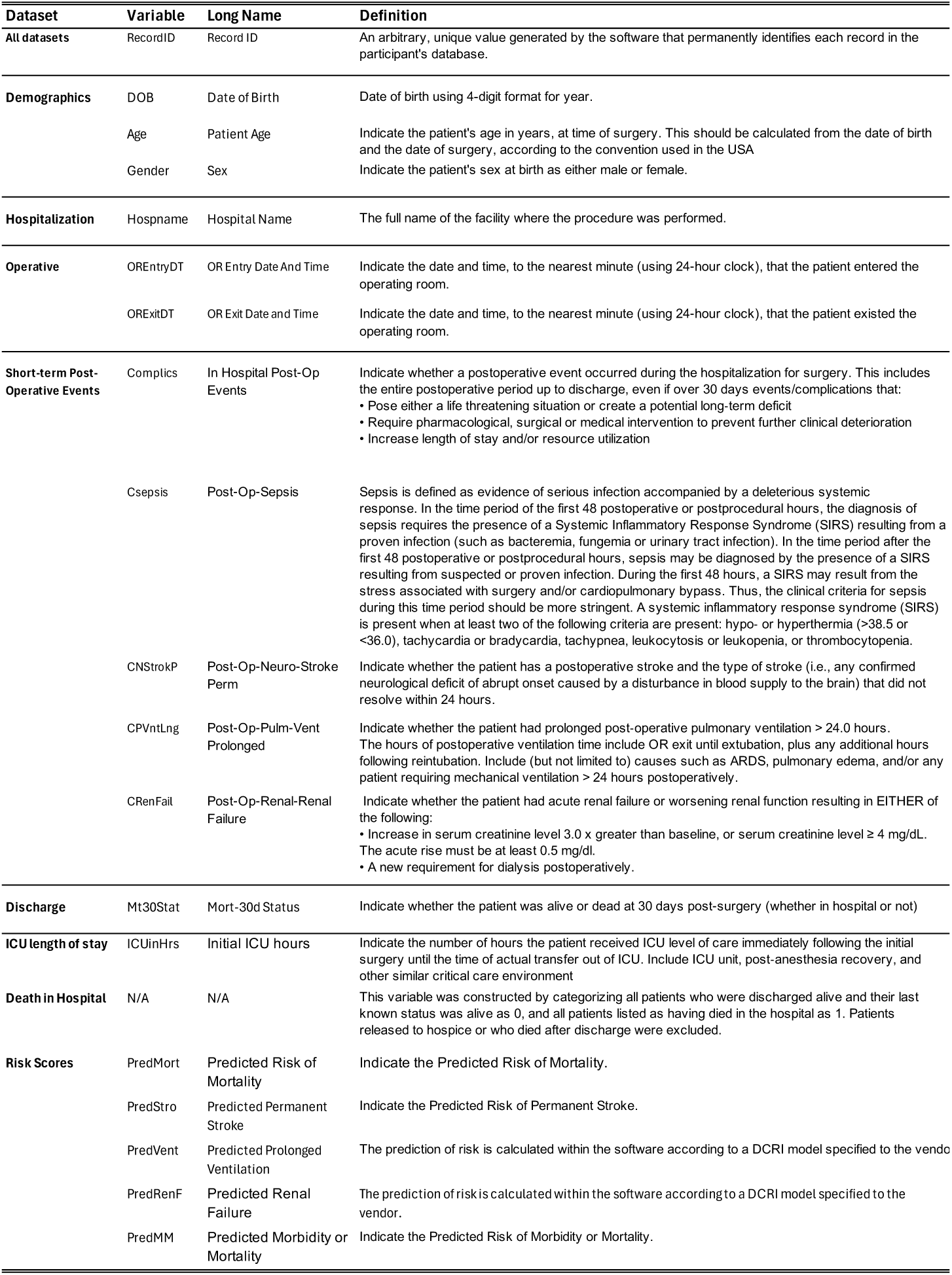
**Definition of clinical outcomes and risk scores used in pre-operative and post­operative analyses.** This table lists clinical outcomes, recovery measures, and Society of Thoracic Surgeons (STS) risk scores used to evaluate pre-operative biological age and early post-operative biological age trajectories. Outcomes include in-hospital stroke, renal failure, prolonged ventilation, sepsis, composite complications, ICU length of stay, and mortality during hospitalization or within 30 days of surgery, as defined in the STS Adult Cardiac Surgeons Database (ACSD) Training Manual. STS risk scores represent model-based predictions derived from pre-operative clinical characteristics, comorbidities, physiologic measures, and procedural factors. Variable names and definitions correspond to STS ACSD conventions.

**Table S2.** **Analytes, panel groups, and measurement numbers in EHR data after two-pass search.**

| **Biomarker** | **Panel Name** | **Number of**<br>**measurements** | **Unique Individuals** |
| --- | --- | --- | --- |
| blood_urea_nitrogen | BASIC METABOLIC PANEL | 136764 | 4313 |
| creatinine | BASIC METABOLIC PANEL | 136597 | 4313 |
| calcium_level | BASIC METABOLIC PANEL | 136568 | 4313 |
| sodium_level | BASIC METABOLIC PANEL | 136442 | 4313 |
| chloride_level | BASIC METABOLIC PANEL | 136405 | 4313 |
| potassium_level | BASIC METABOLIC PANEL | 136318 | 4313 |
| glucose | BASIC METABOLIC PANEL | 132455 | 4313 |
| mean_cell_volume | CBC PLATELET W/O DIFFERENTIAL | 129159 | 4313 |
| mean_cell_hemoglobin | CBC PLATELET W/O DIFFERENTIAL | 129145 | 4313 |
| mean_cell_hemoglobin_concentration | CBC PLATELET W/O DIFFERENTIAL | 129145 | 4313 |
| hematocrit | CBC PLATELET W/O DIFFERENTIAL | 129070 | 4313 |
| white_blood_cell | CBC PLATELET W/O DIFFERENTIAL | 129001 | 4313 |
| red_blood_cell_count | CBC PLATELET W/O DIFFERENTIAL | 128995 | 4313 |
| red_cell_distribution_width | CBC PLATELET W/O DIFFERENTIAL | 128856 | 4313 |
| mean_platelet_volume | CBC PLATELET W/O DIFFERENTIAL | 118555 | 4313 |
| protein_total | HEPATIC FUNCTION PANEL | 95199 | 4313 |
| albumin | HEPATIC FUNCTION PANEL | 94789 | 4313 |
| alkaline_phosphatase | HEPATIC FUNCTION PANEL | 94694 | 4313 |
| aspartate_aminotransferase | HEPATIC FUNCTION PANEL | 94679 | 4313 |
| bilirubin_total | HEPATIC FUNCTION PANEL | 94589 | 4313 |
| globulin | HEPATIC FUNCTION PANEL | 92764 | 4312 |
| alanine_aminotransferase | HEPATIC FUNCTION PANEL | 91384 | 4310 |

**Table S3.** **Analytes, skewness in EHR, and transformation applied in EHR and NHANES prior to model fitting.**

| Biomarker | Skewness | Transformation applied |
| --- | --- | --- |
| alanine_aminotransferase | Skewness = 19.21 → Log recommended | log |
| albumin | Skewness = 7.7 → Log recommended | log |
| alkaline_phosphatase | Skewness = 6.55 → Log recommended | log |
| aspartate_aminotransferase | Skewness = 23.97 → Log recommended | log |
| bilirubin_total | Skewness = 9.79 → Log recommended | log1p |
| blood_urea_nitrogen | Skewness = 2.11 → Log recommended | log |
| calcium_level | Skewness = 0.15 → No log needed | none |
| chloride_level | Skewness = -0.28 → No log needed | none |
| creatinine | Skewness = 3.53 → Log recommended | log |
| globulin | Skewness = 1.07 → Log recommended | log |
| glucose | Skewness = 3.99 → Log recommended | log |
| hematocrit | Skewness = 0.71 → Consider log | log |
| mean_cell_hemoglobin | Skewness = -0.7 → No log needed | none |
| mean_cell_hemoglobin_concentration | Skewness = -0.15 → No log needed | none |
| mean_cell_volume | Skewness = -0.28 → No log needed | none |
| mean_platelet_volume | Skewness = 0.32 → No log needed | none |
| potassium_level | Skewness = 1.07 → Log recommended | log |
| protein_total | Skewness = 6.71 → Log recommended | log |
| red_blood_cell_count | Skewness = 0.74 → Consider log | log |
| red_cell_distribution_width | Skewness = 1.39 → Log recommended | log |
| sodium_level | Skewness = -0.09 → No log needed | none |

**Table S4.**
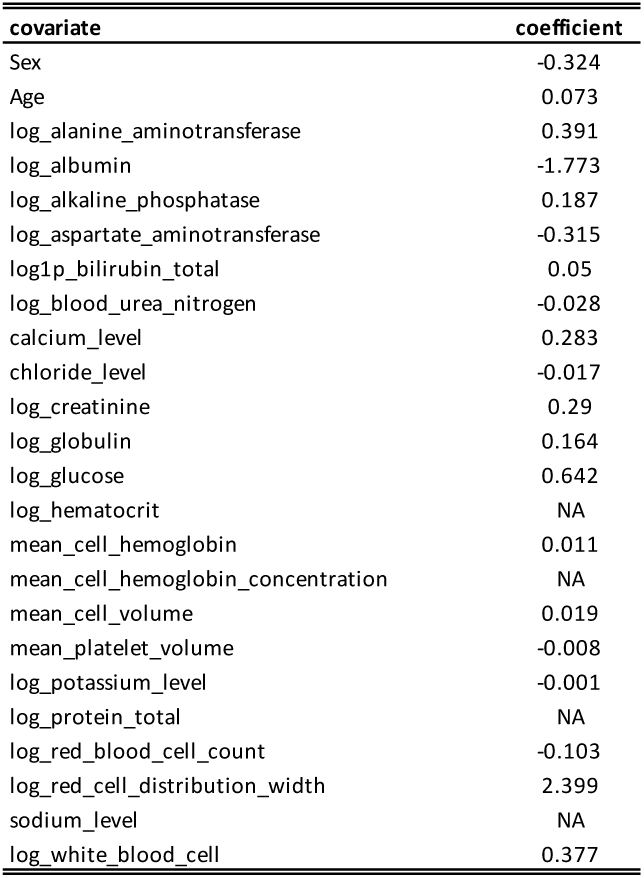
**Coefficients estimated for elastic net Cox regression of mortality in NHANES III with age, sex, and 22 analytes selected from panels in the New York Presbyterian cardiothoracic surgery ward electronic health records.** Coefficients with NA were not selected in the minimum lambda after 10-fold cross-validation.

**Table S5.** **Coefficients from sex-specific Gompertz proportional hazards model relating mortality risk to chronological age and the 18 analytes selected using elastic net Cox regression in NHANES Wave III.**

| covariate | Men | Women |
| --- | --- | --- |
| shape | 0.006 | 0.007 |
| rate | -24.174 | -19.841 |
| age | 0.068 | 0.079 |
| log_alanine_aminotransferase | 0.4 | 0.441 |
| log_albumin | -1.269 | -2.11 |
| log_alkaline_phosphatase | 0.134 | 0.292 |
| log_aspartate_aminotransferase | -0.441 | -0.263 |
| log1p_bilirubin_total | 0.208 | 0.231 |
| log_blood_urea_nitrogen | -0.13 | -0.027 |
| calcium_level | 0.347 | 0.196 |
| chloride_level | -0.006 | -0.02 |
| log_creatinine | 0.011 | 0.699 |
| log_globulin | 0.701 | 0.002 |
| log_glucose | 0.714 | 0.536 |
| mean_cell_hemoglobin | -0.05 | 0.042 |
| mean_cell_volume | 0.042 | 0.004 |
| mean_platelet_volume | -0.017 | -0.026 |
| log_potassium_level | -0.278 | -0.029 |
| log_red_blood_cell_count | -0.925 | 0.324 |
| log_red_cell_distribution_width | 2.716 | 1.563 |
| log_white_blood_cell | 0.476 | 0.351 |

**Table S6.** **Associations of PhenoAge and EHR BioAge measures of biological aging with mortality in NHANES IV.** Coefficients in the table are hazard ratios estimated from Cox proportional hazard regressions. Both PhenoAge and EHR BioAge measures were differenced from chronological age for analysis (i.e. values = BA-CA). These differenced values were then standardized to have M=0, SD=1 separately for men and women within the analysis sample so that effect-sizes are denominated in terms of a sex-specific 1 SD unit increase in biological age advancement. Models included covariates for chronological age and sex. The original Levine Phenotypic Age algorithm was projected onto data from NHANES 1999-2010 and 2015-2018 only because the intervening waves did not include CRP measurements. EHR biological age samples were available for other waves but were restricted to the subset that had PhenoAge for direct comparison.

|  | PhenoAge | EHR Biol. Age |
| --- | --- | --- |
|  | Hazard Ratio (95% CI) |  |
| **Full Sample** |  |  |
| n | 33251 | 33251 |
| BioAge | 1.49 (1.46, 1.52) | 1.45 (1.41, 1.48) |
|  | **Stratified by Gender** |  |
| **Men** |  |  |
| n | 16026 | 16026 |
| BioAge | 1.46 (1.42, 1.5) | 1.45 (1.41, 1.5) |
| **Women** |  |  |
| n | 17225 | 17225 |
| BioAge | 1.55 (1.5, 1.6) | 1.44 (1.38, 1.5) |
|  | **Stratified by Race** |  |
| **White** |  |  |
| n | 15012 | 15012 |
| BioAge | 1.52 (1.48, 1.57) | 1.46 (1.41, 1.51) |
| **Black** |  |  |
| n | 6493 | 6493 |
| BioAge | 1.4 (1.33, 1.47) | 1.41 (1.33, 1.5) |
| **Other** |  |  |
| n | 11746 | 11746 |
| BioAge | 1.52 (1.45, 1.6) | 1.47 (1.39, 1.56) |
|  | **People Aged 65 and Younger** |  |
| **Aged 65 and Younger** |  |  |
| n | 25496 | 25496 |
| BioAge | 1.63 (1.57, 1.69) | 1.65 (1.57, 1.72) |

**Table S7.** Correlations between average pre-operative chronological age and pre­operative biological age for men and women grouped by pre-operative time window (average of all preoperative measurements, measurements within 1 year, measurements within 6 months, and measurements within 3 months).

| Pre-surgery time window | Sex | Correlation | r2 | n |
| --- | --- | --- | --- | --- |
| All presurgery | Female | 0.833 | 0.694 | 1054 |
| All presurgery | Male | 0.814 | 0.663 | 2427 |
| 1y presurgery | Female | 0.828 | 0.686 | 1020 |
| 1y presurgery | Male | 0.811 | 0.657 | 2364 |
| 6m presurgery | Female | 0.828 | 0.685 | 1009 |
| 6m presurgery | Male | 0.808 | 0.653 | 2334 |
| 3m presurgery | Female | 0.829 | 0.686 | 996 |
| 3m presurgery | Male | 0.808 | 0.653 | 2306 |

**Table S8.** **Sociodemographic and clinical variables for all patients in the New York Presbyterian Hospital cardiothoracic surgery record system with laboratory panels for pre-operative (left) and post-operative (right) biological age after exclusion for implausible follow-up dates, readmission to the ICU after discharge, and appropriate data within the 48h post-operative period.**

| Characteristic | N | Lab Sample - Pre<br>N = 3,343<sup>†</sup> | Lab Sample - Post<br>N = 3,991<sup>†</sup> |
| --- | --- | --- | --- |
| Age at Surgery (years) | 7,334 | 63.6 (12.1) | 63.5 (12.0) |
| Sex | 7,334 |  |  |
| Female |  | 1,009 (30.2%) | 1,191 (29.8%) |
| Male |  | 2,334 (69.8%) | 2,800 (70.2%) |
| Race | 7,334 |  |  |
| Asian |  | 101 (3.0%) | 123 (3.1%) |
| Black |  | 305 (9.1%) | 337 (8.4%) |
| Caucasian |  | 2,067 (61.8%) | 2,539 (63.6%) |
| Missing/Undocumented |  | 444 (13.3%) | 502 (12.6%) |
| NatAm_Pacif_Other |  | 426 (12.7%) | 490 (12.3%) |
| Procedure | 7,334 |  |  |
| AV Replacement |  | 330 (9.9%) | 400 (10.0%) |
| AV Replacement + CAB |  | 142 (4.2%) | 165 (4.1%) |
| AV Replacement + MV Replacement |  | 87 (2.6%) | 90 (2.3%) |
| CAB Only |  | 1,306 (39.1%) | 1,527 (38.3%) |
| MV Repair |  | 189 (5.7%) | 275 (6.9%) |
| MV Repair + CAB |  | 33 (1.0%) | 39 (1.0%) |
| MV Replacement + CAB |  | 52 (1.6%) | 56 (1.4%) |
| MV Replacement Only |  | 189 (5.7%) | 211 (5.3%) |
| Other |  | 1,015 (30.4%) | 1,228 (30.8%) |
| Urgency | 7,332 |  |  |
| Full conventional sternotomy |  | 3,073 (92.0%) | 3,605 (90.4%) |
| Limited (mini) thoracotomy, right |  | 1 (0.0%) | 1 (0.0%) |
| Partial sternotomy |  | 94 (2.8%) | 131 (3.3%) |
| Percutaneous |  | 21 (0.6%) | 15 (0.4%) |
| Port access |  | 41 (1.2%) | 105 (2.6%) |
| Right thoracotomy |  | 2 (0.1%) | 2 (0.1%) |
| Sub-xiphoid |  | 4 (0.1%) | 10 (0.3%) |
| Thoracoabdominal incision |  | 14 (0.4%) | 12 (0.3%) |
| Thoracotomy |  | 92 (2.8%) | 109 (2.7%) |
| Surgical Status | 7,332 |  |  |
| Elective |  | 1,812 (54.2%) | 2,439 (61.1%) |
| Emergent |  | 75 (2.2%) | 80 (2.0%) |
| Emergent Salvage |  | 8 (0.2%) | 9 (0.2%) |
| Urgent |  | 1,447 (43.3%) | 1,462 (36.6%) |
| 30-day Mortality | 7,334 |  |  |
| Alive |  | 3,282 (98.2%) | 3,939 (98.7%) |
| Dead |  | 61 (1.8%) | 52 (1.3%) |
| Stroke | 3,453 |  |  |
| No |  | 1,556 (96.2%) | 1,774 (96.6%) |
| Yes |  | 61 (3.8%) | 62 (3.4%) |
| Yes, embolic |  | 0 (0.0%) | 0 (0.0%) |
| Yes, hemorrhagic |  | 0 (0.0%) | 0 (0.0%) |
| Yes, undetermined type |  | 0 (0.0%) | 0 (0.0%) |
| Prolonged Ventilation | 3,453 | 557 (34.4%) | 537 (29.2%) |
| Renal Failure | 3,453 | 129 (8.0%) | 116 (6.3%) |
| Sepsis | 3,453 | 59 (3.6%) | 38 (2.1%) |
| Complications | 7,333 | 1,617 (48.4%) | 1,836 (46.0%) |

<sup>†</sup> Mean (SD); n (%)

**Table S9.** **Risk ratios and 95% confidence intervals for associations between average 6-month pre-operative biological age and six surgery outcomes (mortality status at 30d post-surgery, stroke, renal failure, prolonged ventilation, sepsis, or other complications), stratified by sex.**

| Outcome | Sex | N | events | Risk ratio | Lower 95% CI | Upper 95% CI | P-value |
| --- | --- | --- | --- | --- | --- | --- | --- |
| Mortality status (30d post-surgery) | Men | 2334 | 37 | 1.62 | 1.39 | 1.88 | <0.001 |
|  | Women | 1009 | 24 | 1.59 | 1.39 | 1.82 | <0.001 |
| Stroke | Men | 1095 | 41 | 1.18 | 0.98 | 1.43 | 0.088 |
|  | Women | 522 | 20 | 1.36 | 1.13 | 1.63 | 0.001 |
| Renal failure | Men | 1095 | 76 | 1.25 | 1.10 | 1.41 | <0.001 |
|  | Women | 522 | 53 | 1.22 | 1.12 | 1.33 | <0.001 |
| Prolonged ventilation | Men | 1095 | 351 | 1.30 | 1.25 | 1.36 | <0.001 |
|  | Women | 522 | 206 | 1.18 | 1.14 | 1.23 | <0.001 |
| Sepsis | Men | 1095 | 39 | 1.69 | 1.45 | 1.96 | <0.001 |
|  | Women | 522 | 20 | 1.44 | 1.22 | 1.71 | <0.001 |
| Complications | Men | 2334 | 1095 | 1.13 | 1.10 | 1.16 | <0.001 |
|  | Women | 1008 | 522 | 1.10 | 1.07 | 1.13 | <0.001 |

**Table S10.** **Comparison of sociodemographic and clinical variables for patients with pre­operative laboratory panels used to calculate EHR biological age and the nested subset of patients who also had Society for Thoracic (STS) risk scores.**

| Characteristic | N | Lab Sample - Pre<br>N = 3,343<sup>1</sup> | STS Sample - Pre<br>N = 2,322<sup>2</sup> | p-value<sup>2</sup> |
| --- | --- | --- | --- | --- |
| Age at Surgery (years) | 5,665 | 63.6 (12.1) | 65.1 (10.6) | **<0.001** |
| Sex | 5,665 |  |  | 0.093 |
| Female |  | 1,009 (30.2%) | 652 (28.1%) |  |
| Male |  | 2,334 (69.8%) | 1,670 (71.9%) |  |
| Race | 5,665 |  |  | 0.2 |
| Asian |  | 101 (3.0%) | 71 (3.1%) |  |
| Black |  | 305 (9.1%) | 172 (7.4%) |  |
| Caucasian |  | 2,067 (61.8%) | 1,457 (62.7%) |  |
| Missing/Undocumented |  | 444 (13.3%) | 300 (12.9%) |  |
| NatAm_Pacif_Other |  | 426 (12.7%) | 322 (13.9%) |  |
| Procedure | 5,665 |  |  | **<0.001** |
| AV Replacement |  | 330 (9.9%) | 343 (14.8%) |  |
| AV Replacement + CAB |  | 142 (4.2%) | 146 (6.3%) |  |
| AV Replacement + MV Replacement |  | 87 (2.6%) | 0 (0.0%) |  |
| CAB Only |  | 1,306 (39.1%) | 1,350 (58.1%) |  |
| MV Repair |  | 189 (5.7%) | 202 (8.7%) |  |
| MV Repair + CAB |  | 33 (1.0%) | 34 (1.5%) |  |
| MV Replacement + CAB |  | 52 (1.6%) | 53 (2.3%) |  |
| MV Replacement Only |  | 189 (5.7%) | 194 (8.4%) |  |
| Other |  | 1,015 (30.4%) | 0 (0.0%) |  |
| Urgency | 5,663 |  |  | **<0.001** |
| Full conventional sternotomy |  | 3,073 (92.0%) | 2,135 (92.0%) |  |
| Limited (mini) thoracotomy, right |  | 1 (0.0%) | 1 (0.0%) |  |
| Partial sternotomy |  | 94 (2.8%) | 59 (2.5%) |  |
| Percutaneous |  | 21 (0.6%) | 0 (0.0%) |  |
| Port access |  | 41 (1.2%) | 45 (1.9%) |  |
| Right thoracotomy |  | 2 (0.1%) | 0 (0.0%) |  |
| Sub-xiphoid |  | 4 (0.1%) | 4 (0.2%) |  |
| Thoracoabdominal incision |  | 14 (0.4%) | 0 (0.0%) |  |
| Thoracotomy |  | 92 (2.8%) | 77 (3.3%) |  |
| Surgical Status | 5,663 |  |  | **0.001** |
| Elective |  | 1,812 (54.2%) | 1,293 (55.7%) |  |
| Emergent |  | 75 (2.2%) | 22 (0.9%) |  |
| Emergent Salvage |  | 8 (0.2%) | 2 (0.1%) |  |
| Urgent |  | 1,447 (43.3%) | 1,004 (43.3%) |  |
| 30-day Mortality | 5,665 |  |  | 0.14 |
| Alive |  | 3,282 (98.2%) | 2,292 (98.7%) |  |
| Dead |  | 61 (1.8%) | 30 (1.3%) |  |
| Stroke | 2,630 |  |  | 0.8 |
| No |  | 1,556 (96.2%) | 978 (96.5%) |  |
| Yes |  | 61 (3.8%) | 35 (3.5%) |  |
| Yes, embolic |  | 0 (0.0%) | 0 (0.0%) |  |
| Yes, hemorrhagic |  | 0 (0.0%) | 0 (0.0%) |  |
| Yes, undetermined type |  | 0 (0.0%) | 0 (0.0%) |  |
| Prolonged Ventilation | 2,630 | 557 (34.4%) | 243 (24.0%) | **<0.001** |
| Renal Failure | 2,630 | 129 (8.0%) | 53 (5.2%) | **0.009** |
| Sepsis | 2,630 | 59 (3.6%) | 29 (2.9%) | 0.3 |
| Complications | 5,664 | 1,617 (48.4%) | 1,013 (43.6%) | **<0.001** |

<sup>1</sup> Mean (SD); n (%) <sup>2</sup> Welch Two Sample t-test; Pearson's Chi-squared test

**Table S11.** **Risk ratios and 95% confidence intervals for associations between average 6-month pre-operative EHR biological age and six surgery outcomes (mortality status at 30d post-surgery, stroke, renal failure, prolonged ventilation, sepsis, or other complications).** Estimates come from models that include EHR biological age only in the full sample with EHR biological age and outcome data, EHR biological age only in the sample with biological age, outcome data, and the corresponding STS-predictor, and the EHR biological age estimate in a joint model includes both EHR biological age and the corresponding STS-predictor.

| Outcome | N | events | Model | Risk ratio | Lower 95% CI | Upper 95% CI | P-value |
| --- | --- | --- | --- | --- | --- | --- | --- |
| Mortality status (30d post-surgery) | 3343 | 61 | Biol. Age (Full sample) | 1.61 | 1.46 | 1.78 | <0.001 |
|  | 2241 | 30 | Biol. Age (Overlap subset) | 1.71 | 1.48 | 1.97 | <0.001 |
|  | - | - | Biol. Age (Joint model w/ PredMort) | 1.14 | 0.93 | 1.40 | 0.206 |
| Stroke | 1617 | 61 | Biol. Age (Full sample) | 1.26 | 1.10 | 1.44 | <0.001 |
|  | 981 | 35 | Biol. Age (Overlap subset) | 1.24 | 1.03 | 1.48 | 0.021 |
|  | - | - | Biol. Age (Joint model w/ PredStro) | 1.06 | 0.86 | 1.31 | 0.598 |
| Renal failure | 1617 | 129 | Biol. Age (Full sample) | 1.24 | 1.15 | 1.33 | <0.001 |
|  | 932 | 51 | Biol. Age (Overlap subset) | 1.67 | 1.44 | 1.95 | <0.001 |
|  | - | - | Biol. Age (Joint model w/ PredRenf) | 1.12 | 0.90 | 1.40 | 0.313 |
| Prolonged ventilation | 1617 | 557 | Biol. Age (Full sample) | 1.24 | 1.20 | 1.28 | <0.001 |
|  | 981 | 235 | Biol. Age (Overlap subset) | 1.30 | 1.23 | 1.37 | <0.001 |
|  | - | - | Biol. Age (Joint model w/ PredVent) | 1.06 | 1.00 | 1.14 | 0.063 |
| Sepsis | 1617 | 59 | Biol. Age (Full sample) | 1.54 | 1.38 | 1.72 | <0.001 |
|  | 981 | 29 | Biol. Age (Overlap subset) | 1.67 | 1.41 | 1.98 | <0.001 |
|  | - | - | Biol. Age (Joint model w/ PredMM) | 1.29 | 1.04 | 1.61 | 0.023 |
| Complications | 3342 | 1617 | Biol. Age (Full sample) | 1.12 | 1.10 | 1.14 | <0.001 |
|  | 2241 | 981 | Biol. Age (Overlap subset) | 1.11 | 1.08 | 1.14 | <0.001 |
|  | - | - | Biol. Age (Joint model w/ PredMM) | 1.03 | 1.00 | 1.07 | 0.089 |

**Table S12.** **Receiver operating characteristic area-under-the-curve (AUC) for different models predicting six adverse outcomes following cardiothoracic surgery: mortality, stroke, renal failure, prolonged ventilation, sepsis, and composite complications.** For each outcome, we show AUC for in the subset of patients with EHR biological age and outcome data (‘full sample’) or the subset with both EHR biological age and the corresponding STS predicted risk score available (‘overlap sample’). Estimated AUCs and 95% confidence intervals were estimated using the pROC package in R^29^ for models with raw 6-month average pre­operative EHR biological age in the full or overlap sample, the log-transformed STS risk score for the overlap sample, and the AUC from a combined model including log(STS score), raw EHR biological age, age, age², and sex.

| Outcome | Sample | N | N events | Biol. Age AUC<br>(95% CI) | STS predictor<br>AUC [95% CI] | Combined AUC<br>[95% CI] |
| --- | --- | --- | --- | --- | --- | --- |
| Mortality status | Pre-surgery Biol. Age (full sample) | 2241 | 61 | 0.73 [0.66,0.81] | - | - |
|  | Intersection (overlap sample) | 2241 | 30 | 0.71 [0.59,0.82] | 0.83 [0.74,0.91] | 0.82 [0.74,0.91] |
| Stroke | Pre-surgery Biol. Age (full sample) | 1617 | 61 | 0.55 [0.46,0.63] | - | - |
|  | Intersection (overlap sample) | 981 | 35 | 0.58 [0.47,0.68] | 0.69 [0.60,0.78] | 0.69 [0.60,0.79] |
| Renal failure | Pre-surgery Biol. Age (full sample) | 1617 | 129 | 0.57 [0.52,0.62] | - | - |
|  | Intersection (overlap sample) | 932 | 51 | 0.66 [0.59,0.74] | 0.82 [0.75,0.88] | 0.82 [0.75,0.88] |
| Prolonged ventilation | Pre-surgery Biol. Age (full sample) | 1617 | 557 | 0.58 [0.55,0.61] | - | - |
|  | Intersection (overlap sample) | 981 | 235 | 0.59 [0.55,0.64] | 0.75 [0.72,0.79] | 0.76 [0.72,0.79] |
| Sepsis | Pre-surgery Biol. Age (full sample) | 1617 | 59 | 0.72 [0.66,0.79] | - | - |
|  | Intersection (overlap sample) | 981 | 29 | 0.77 [0.69,0.86] | 0.81 [0.74,0.89] | 0.83 [0.76,0.91] |
| Complications | Pre-surgery Biol. Age (full sample) | 3342 | 1617 | 0.63 [0.61,0.64] | - | - |
|  | Intersection (overlap sample) | 2241 | 981 | 0.62 [0.60,0.65] | 0.63 [0.61,0.66] | 0.65 [0.62,0.67] |

**Table S13.**
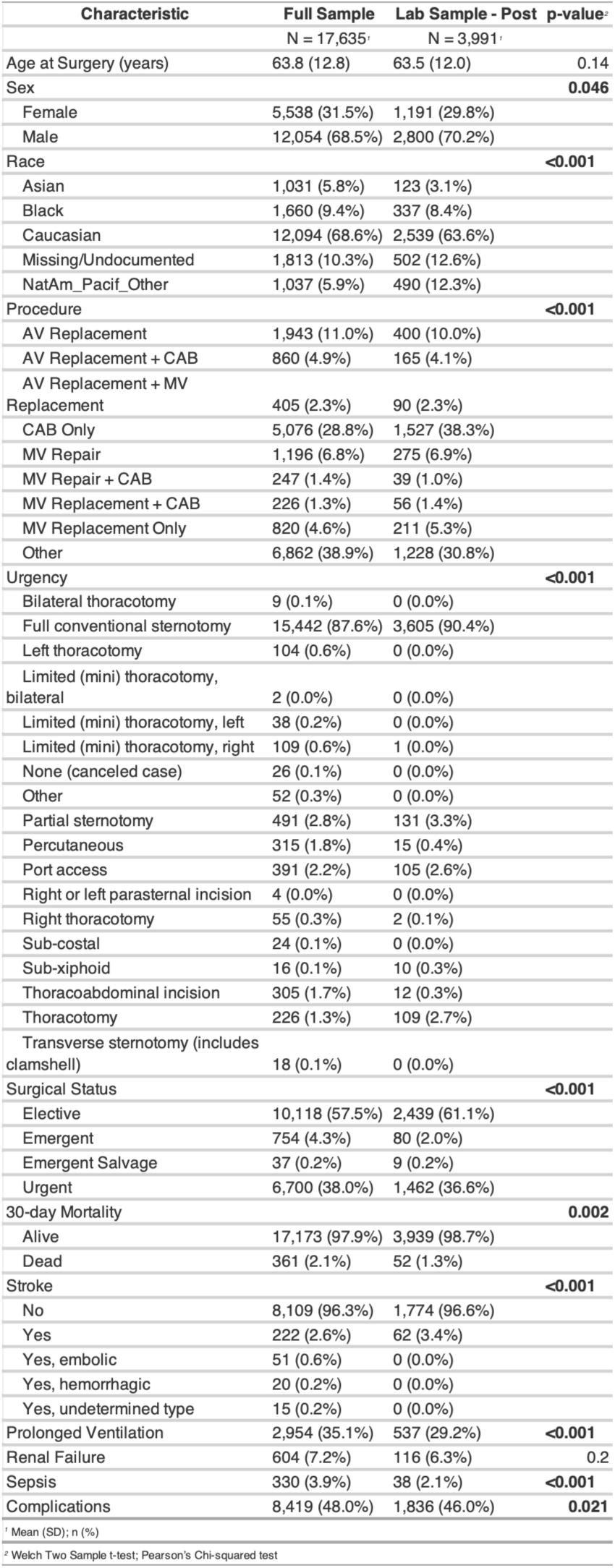
**Comparison of sociodemographic and clinical variables for all patients in the New York Presbyterian Hospital thoracic surgery record system and the nested subset of patients with post-surgery laboratory panels used to calculate EHR biological age taken within the first 48-hours after surgery.**

**Table S14.** **Association between early post-operative biological age and post-operative outcomes (ICU length of stay, chronic critical Illness, and In-hospital mortality).** Biological age was defined as each individual’s average biological age across the first 48 hours following surgery. Models adjust for relevant demographic and clinical covariates. ICU length of stay (left) was modelled using Cox proportional hazards models to evaluate the association between biological age and time to ICU discharge. Estimates are reported as hazard ratios (HRs) with 95% confidence intervals, obtained by exponentiating the model coefficients. Hazard ratios <1 indicate a lower rate of ICU discharge (i.e., longer ICU stay), whereas hazard ratios >1 indicate a higher rate of ICU discharge. Chronic critical illness (defined as remaining in the ICU for more than 7 days following surgery; middle), and in-hospital mortality (right) were modelled using generalized linear models with a logit link. Estimates are reported as odds ratios (ORs) with 95% confidence intervals, obtained by exponentiating the model coefficients. Odds ratios >1 indicate higher odds of prolonged ICU stay or death during hospital stay.

| *Predictors* | **Cox predicting ICU hours (all patients)** |  |  | **GLM predicting chronic illness (>7d)** |  |  | **GLM predicting In hospital death** |  |  |
| --- | --- | --- | --- | --- | --- | --- | --- | --- | --- |
|  | *Estimates* | *CI* | *p* | *Odds Ratios* | *CI* | *p* | *Odds Ratios* | *CI* | *p* |
| Age | 1.01 | 0.99 – 1.03 | 0.319 | 0.94 | 0.81 – 1.14 | 0.502 | 1 | 0.94 – 1.07 | 0.96 |
| Age Squared/1000 | 0.83 | 0.71 – 0.97 | **0.018** | 2.57 | 0.57 – 9.18 | 0.178 | 1.18 | 0.70 – 1.92 | 0.529 |
| Sex (M) | 0.95 | 0.89 – 1.02 | 0.172 | 0.67 | 0.39 – 1.18 | 0.166 | 0.9 | 0.73 – 1.12 | 0.351 |
| Avg. Biol. Age (first 48h in ICU) | 0.64 | 0.62 – 0.67 | **<0.001** | 3.03 | 2.46 – 3.76 | **<0.001** | 2.41 | 2.19 – 2.66 | **<0.001** |
| Intercept |  |  |  | 0.01 | 0.00 – 0.73 | 0.065 | 0.05 | 0.01 – 0.31 | **0.002** |
| Observations | 3991 |  |  | 3968 |  |  | 3991 |  |  |
| R<sup>2</sup> Nagelkerke | 0.168 |  |  | 0.089 |  |  | 0.114 |  |  |

**Table S15.** **Associations of postoperative EHR BioAge trajectory components with ICU duration, chronic critical illness, and in-hospital mortality across alternative model specifications.** Participant-specific postoperative EHR BioAge intercepts and slopes were estimated from a linear mixed-effects model of repeated EHR BioAge measurements obtained during the first 48 hours after surgery. The mixed-effects model included postoperative time, age at surgery, age squared, and sex as fixed effects and participant-specific random intercepts and slopes. Panel A presents hazard ratios from Cox proportional hazards models of time to live ICU discharge; hazard ratios below 1 indicate a lower rate of discharge and longer ICU stay. Panels B and C present odds ratios from generalized linear models with a binomial distribution and logit link for chronic critical illness, defined as ICU stay exceeding 7 days, and in-hospital mortality, respectively. Each panel presents models containing the postoperative EHR BioAge intercept alone, slope alone, intercept and slope, slope and slope squared, or intercept, slope, and slope squared. All outcome models were adjusted for age at surgery, age squared, and sex. EHR BioAge intercepts and slopes were standardized, and slope squared was calculated as the square of the standardized slope. Estimates are presented as hazard ratios or odds ratios with 95% confidence intervals and p-values. An em dash indicates that a trajectory component was not included in the model.

| Model specification | BioAge intercept |  | BioAge slope |  | BioAge slope<sup>2</sup> |  |
| --- | --- | --- | --- | --- | --- | --- |
|  | Effect estimate (95% CI) | p-value | Effect estimate (95% CI) | p-value | Effect estimate (95% CI) | p-value |
| **Panel A. ICU duration (N = 3,991; hazard ratios for live ICU discharge)** |  |  |  |  |  |  |
| Intercept only | 0.69 (0.67–0.71) | <0.001 | — | — | — | — |
| Slope only | — | — | 1.06 (1.03–1.09) | <0.001 | — | — |
| Intercept + slope | 0.69 (0.66–0.71) | <0.001 | 0.97 (0.94–1.00) | 0.069 | — | — |
| Slope + slope<sup>2</sup> | — | — | 1.06 (1.02–1.09) | 0.001 | 0.90 (0.89–0.92) | <0.001 |
| **Intercept + slope + slope<sup>2</sup>** | **0.70 (0.68–0.72)** | **<0.001** | **0.97 (0.94–1.00)** | **0.068** | **0.93 (0.91–0.95)** | **<0.001** |
| **Panel B. Chronic critical illness (N = 3,991; odds ratios)** |  |  |  |  |  |  |
| Intercept only | 2.24 (2.05–2.46) | <0.001 | — | — | — | — |
| Slope only | — | — | 0.86 (0.78–0.95) | 0.002 | — | — |
| Intercept + slope | 2.31 (2.10–2.55) | <0.001 | 1.12 (1.01–1.23) | 0.028 | — | — |
| Slope + slope<sup>2</sup> | — | — | 0.90 (0.82–0.98) | 0.016 | 1.12 (1.07–1.16) | <0.001 |
| **Intercept + slope + slope<sup>2</sup>** | **2.27 (2.06–2.50)** | **<0.001** | **1.13 (1.02–1.24)** | **0.014** | **1.05 (1.01–1.09)** | **0.024** |
| **Panel C. In-hospital mortality (N = 3,968; odds ratios)** |  |  |  |  |  |  |
| Intercept only | 2.87 (2.34–3.54) | <0.001 | — | — | — | — |
| Slope only | — | — | 0.97 (0.74–1.28) | 0.818 | — | — |
| Intercept + slope | 2.98 (2.42–3.68) | <0.001 | 1.23 (1.00–1.53) | 0.053 | — | — |
| Slope + slope<sup>2</sup> | — | — | 0.95 (0.77–1.16) | 0.617 | 1.15 (1.08–1.22) | <0.001 |
| **Intercept + slope + slope<sup>2</sup>** | **2.83 (2.28–3.54)** | **<0.001** | **1.20 (0.97–1.48)** | **0.084** | **1.07 (0.99–1.15)** | **0.085** |

**Table S16.** **Discrimination of postoperative EHR BioAge trajectory models for chronic critical illness and in-hospital mortality**. Areas under the receiver operating characteristic curve (AUCs) with 95% confidence intervals are presented for generalized linear models with a binomial distribution and logit link predicting chronic critical illness, defined as ICU stay exceeding 7 days (Panel A), and in-hospital mortality (Panel B). The basic model (M1) included age at surgery, age squared, and sex. Subsequent models added the participant-specific postoperative EHR BioAge random intercept (M2), random slope (M3), random intercept and slope (M4), random slope and slope squared (M5), or random intercept, slope, and slope squared (M6). Random intercepts and slopes were estimated from a linear mixed-effects model of repeated EHR BioAge measurements obtained during the first 48 hours after surgery. EHR BioAge intercepts and slopes were standardized, and slope squared was calculated as the square of the standardized slope. AUCs were calculated from model-predicted probabilities in the analytic sample; an AUC of 0.50 indicates chance-level discrimination and an AUC of 1.00 indicates perfect discrimination. Shaded rows identify the full trajectory specification (M6). AUC, area under the receiver operating characteristic curve; CI, confidence interval.

| Model specification | AUC | 95% CI |
| --- | --- | --- |
| **Panel A. Chronic critical illness (>7 days in ICU; N = 3,991; 494 events)** |  |  |
| M1: Basic model (age, age squared, sex) | 0.563 | 0.536–0.590 |
| M2: M1 + random intercept | 0.744 | 0.720–0.768 |
| M3: M1 + random slope | 0.576 | 0.547–0.604 |
| M4: M1 + random intercept + slope | 0.745 | 0.721–0.770 |
| M5: M1 + random slope + slope squared | 0.619 | 0.592–0.645 |
| **M6: M1 + random intercept + slope + slope squared** | **0.749** | **0.725–0.773** |
| **Panel B. In-hospital mortality (N = 3,968; 56 deaths)** |  |  |
| M1: Basic model (age, age squared, sex) | 0.661 | 0.584–0.738 |
| M2: M1 + random intercept | 0.848 | 0.800–0.896 |
| M3: M1 + random slope | 0.667 | 0.592–0.742 |
| M4: M1 + random intercept + slope | 0.845 | 0.796–0.893 |
| M5: M1 + random slope + slope squared | 0.721 | 0.650–0.791 |
| **M6: M1 + random intercept + slope + slope squared** | **0.852** | **0.806–0.898** |

## Supplementary Figures

**Figure S1.**
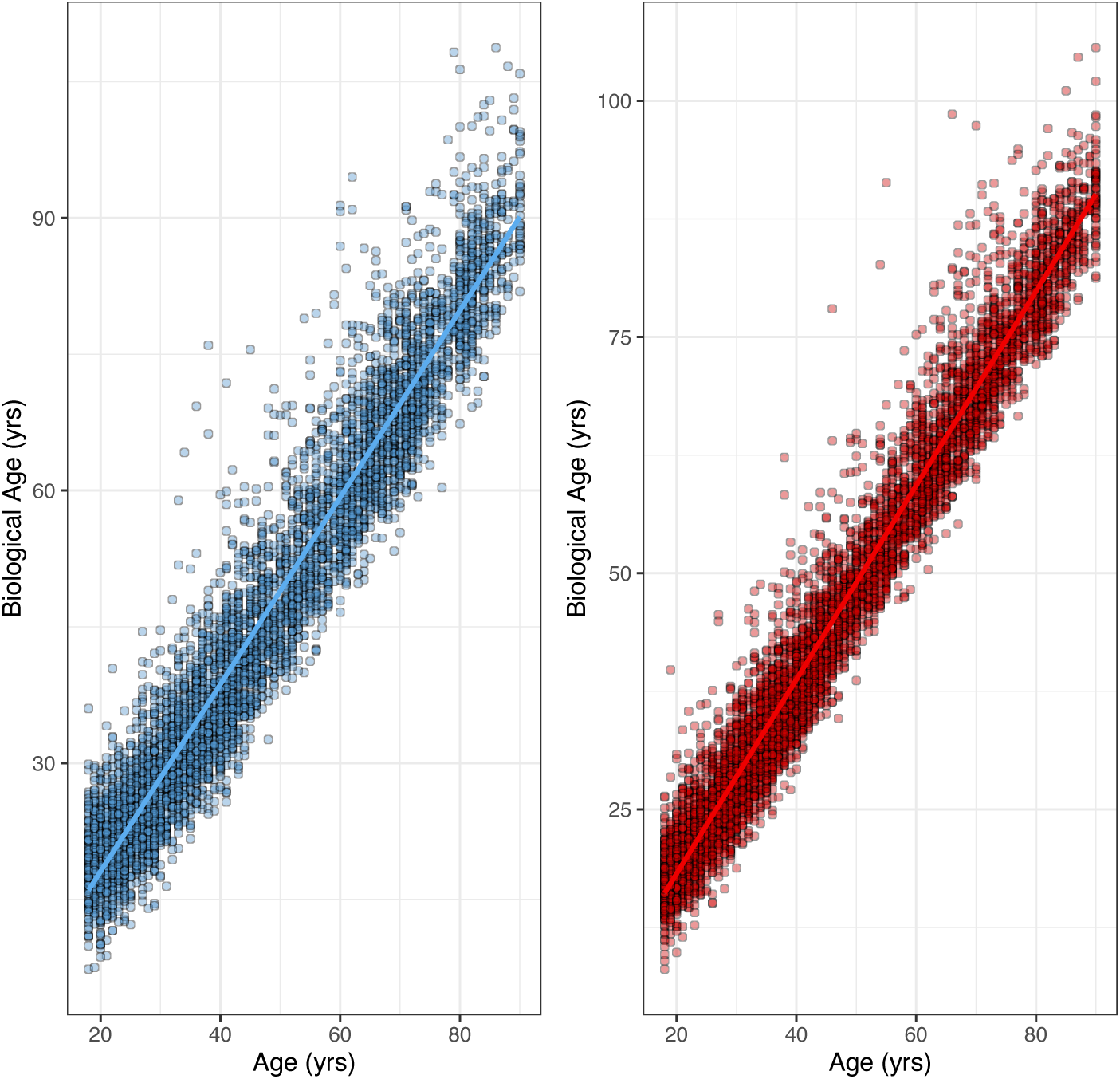
Association between chronological age and estimated biological age in NHANES III training data. Men are plotted on the left in blue, while women are plotted on the right in red. Each point represents an individual. The solid blue (men) and red (women) lines indicate unity, where chronological age and estimated biological age are identical.

**Figure S2.**
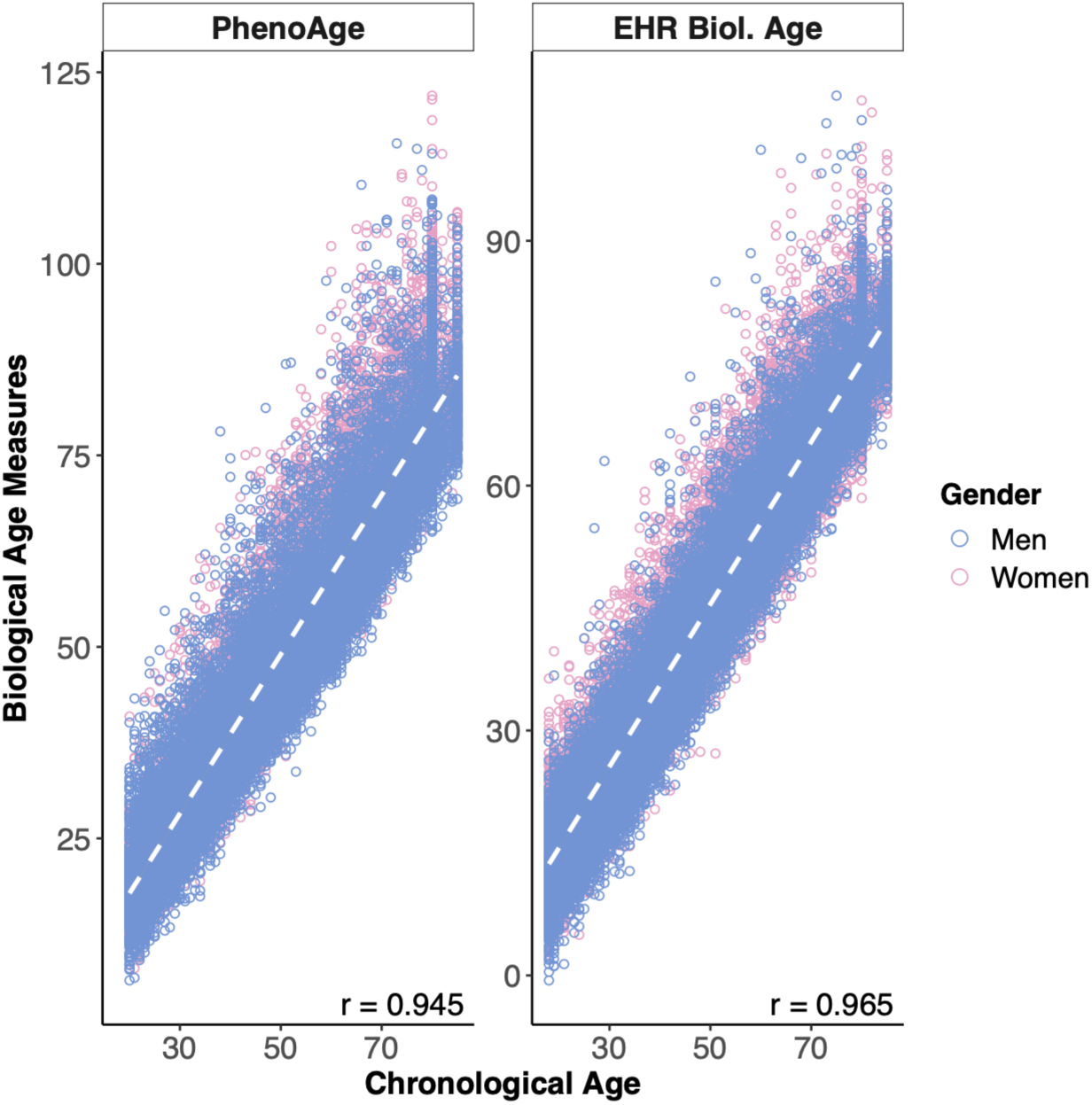
**Association of biological aging measures with chronological age in NHANES IV test data.** The panel on the left shows measures based on the original biomarker set published in <u>Levine 2013 J Geron A</u> and <u>Levine et al. 2018 AGING</u>. The panel on the right shows EHR biological age measure computed with the biomarkers from the EHR data and developed in NHANES III.

**Figure S3.**
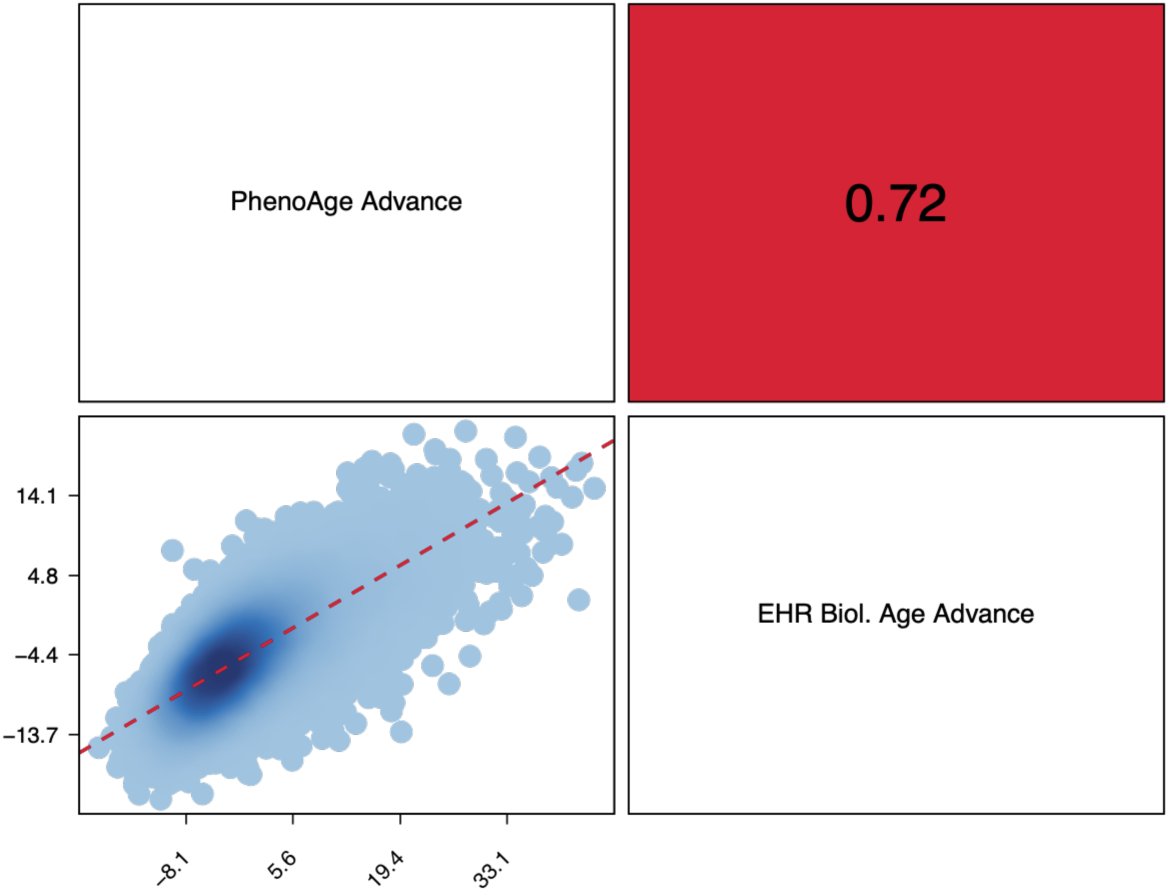
**Correlations among PhenoAge and EHR biological age in NHANES IV test data.** The figure plots associations among Levine’s PhenoAge and EHR PhenoAge, developed in NHANES III and trained in NHANES IV. The cell in the bottom left-hand side shows a scatter plot of PhenoAge and EHR biological age. The cell in the top right-hand side shows the Pearson correlation between PhenoAge and EHR biological age. Measures are differenced from chronological age (i.e. plotted values = BA-CA).

**Figure S4.**
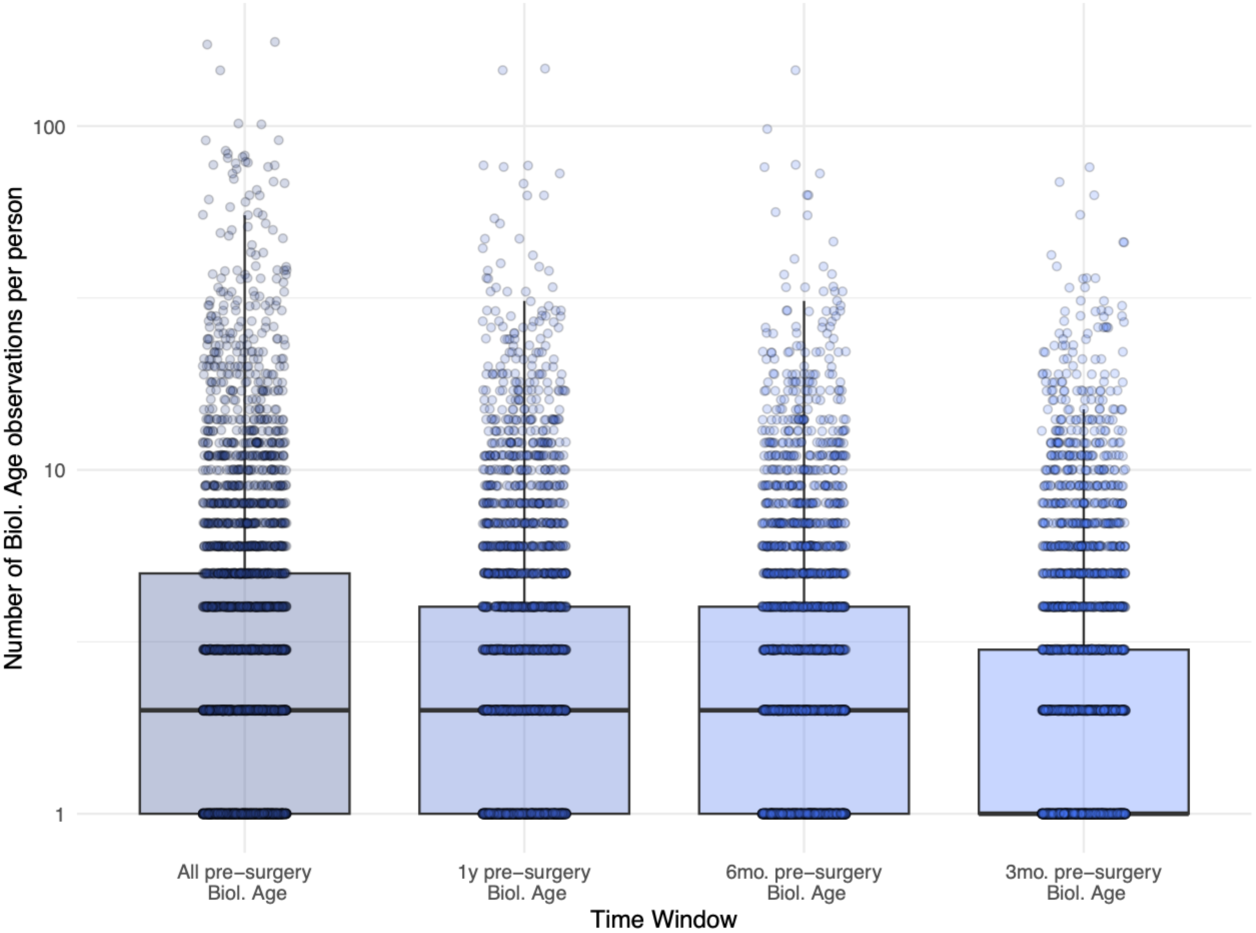
Number of biological age observations per person for four different pre-operative time windows (all pre-operative measures, measures within 1-year, measures within 6-months, and measures within 3-months). The 1^st^ and 3^rd^ quartiles and the median are shown by the lower and upper edges of the box and the horizontal line inside the box, respectively. The whisker shows the points within 1.5x the interquartile range. Median number of biological age measurements for all pre-operative windows were 2 times, except the 3-month pre-operative window where the median number of biological age measurements was once. The X-axis is on a log-scale for legibility.

**Figure S5.**
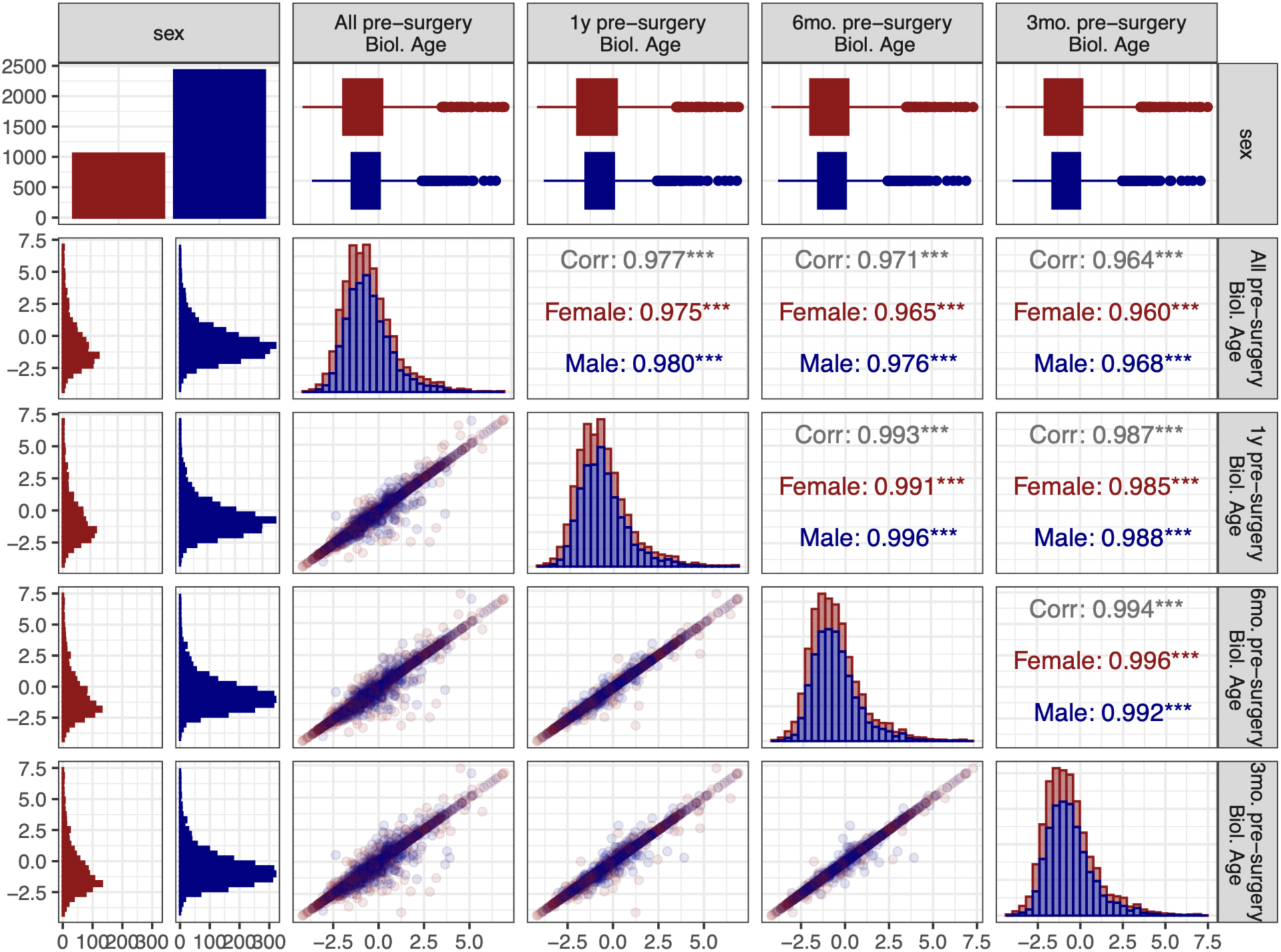
Scatterplots, histograms, and boxplots for average pre-operative biological age calculated in electronic health records using different pre-operative time-windows (all pre­operative measures, measures within 1-year, measures within 6-months, and measures within 3-months). Men are plotted in navy blue, women are plotted in firebrick. Pearson’s r coefficients for both sexes (gray) or each sex separately are shown in the upper diagonal.

**Figure S6.**
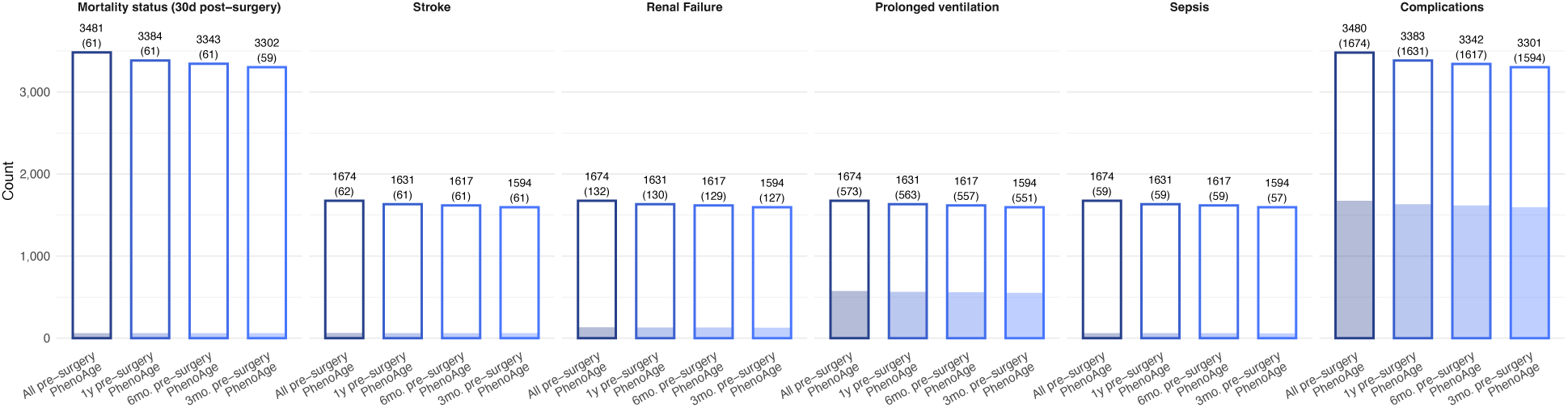
Barchart depicting the number of patients (hollow bar) and events (shaded portion of the bar) for patients with biological age measures during different pre-operative time-windows (all pre-operative measures, measures within 1-year, measures within 6-months, and measures within 3-months). Number of patients (and events) are also shown above each bar.

**Figure S7.**
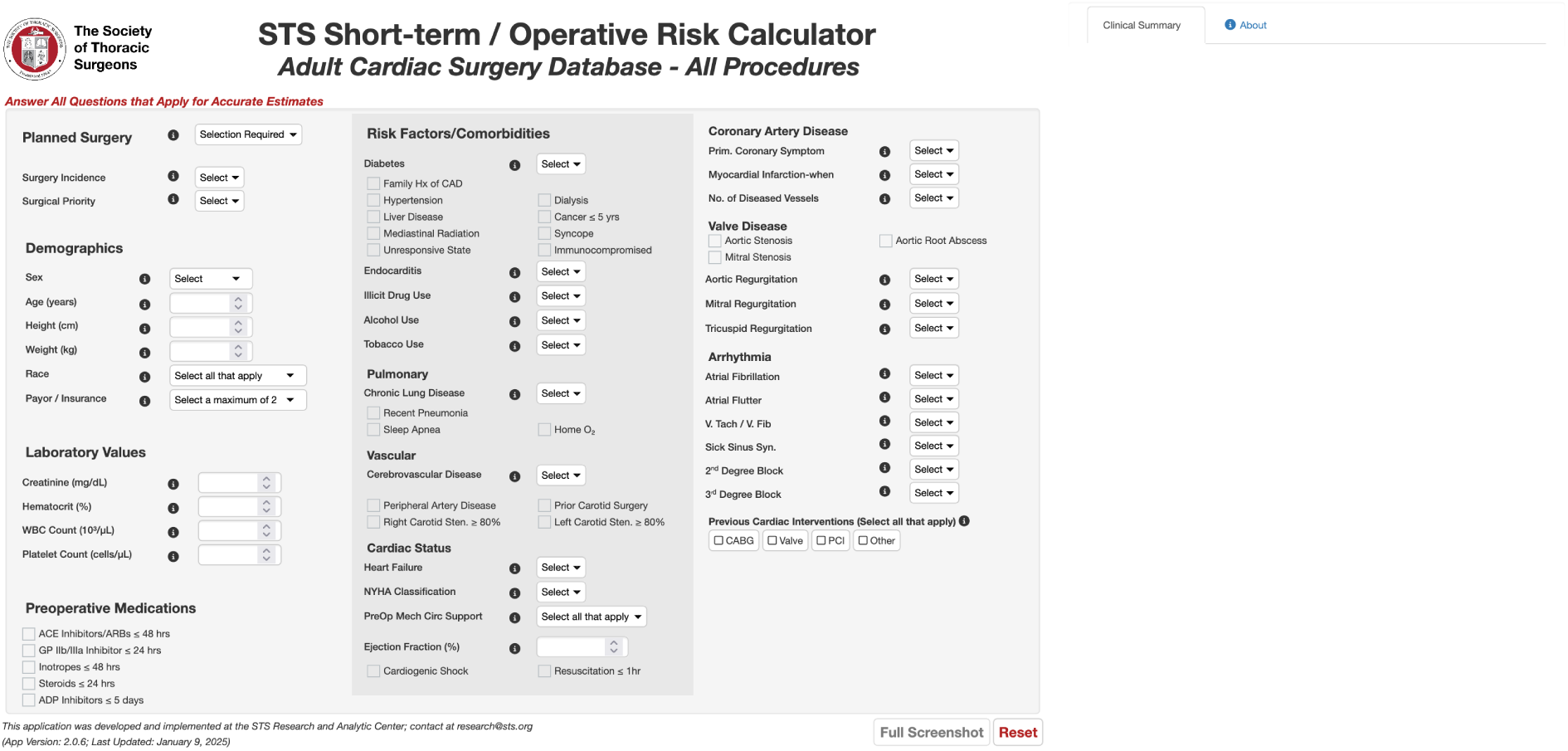
Screenshot of the online risk calculator for the Society of Thoracic Surgeons (STS). The risk predictor takes information about operative priority, demographics, laboratory tests, pre-operative medications, and other risk factors to calculate the risk for a number of post-operational health outcomes. The development of this measure is described in ^22,23^. The online calculator can be accessed at https://www.sts.org/resources/acsd-operative-risk-calculator.

**Figure S8.**
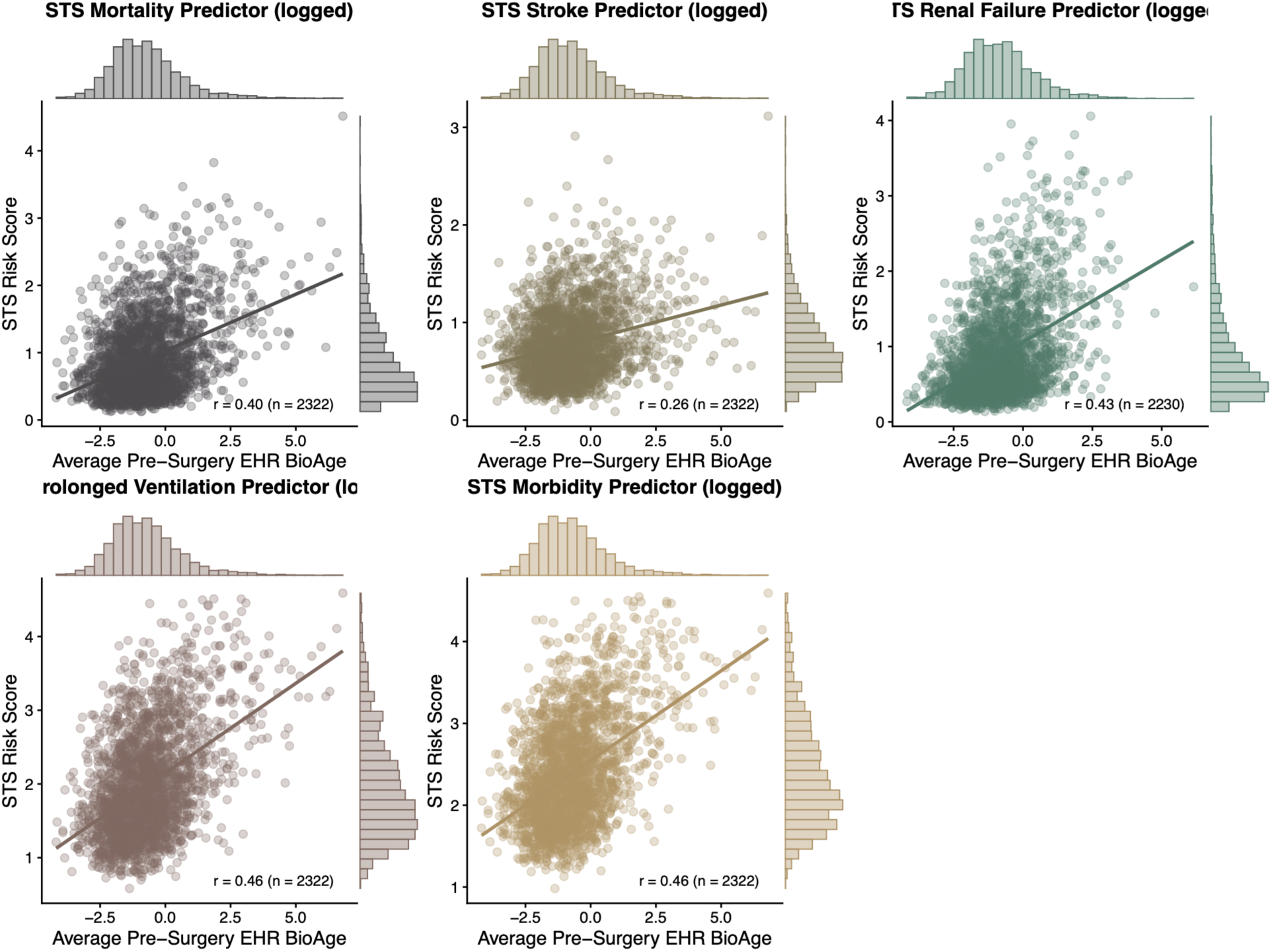
**Association between pre-operative EHR BioAge and STS risk predictors.** Scatterplots show the association between average pre-operative EHR BioAge, scaled in NHANES standard-deviation units, and logged STS risk predictor values for mortality, stroke, renal failure, prolonged ventilation, and morbidity. Points represent individual patients; solid lines show linear regression fits. Marginal histograms display the distribution of EHR BioAge and each STS predictor. Pearson correlation coefficients and analytic sample sizes are shown within each panel.

**Figure S9.**
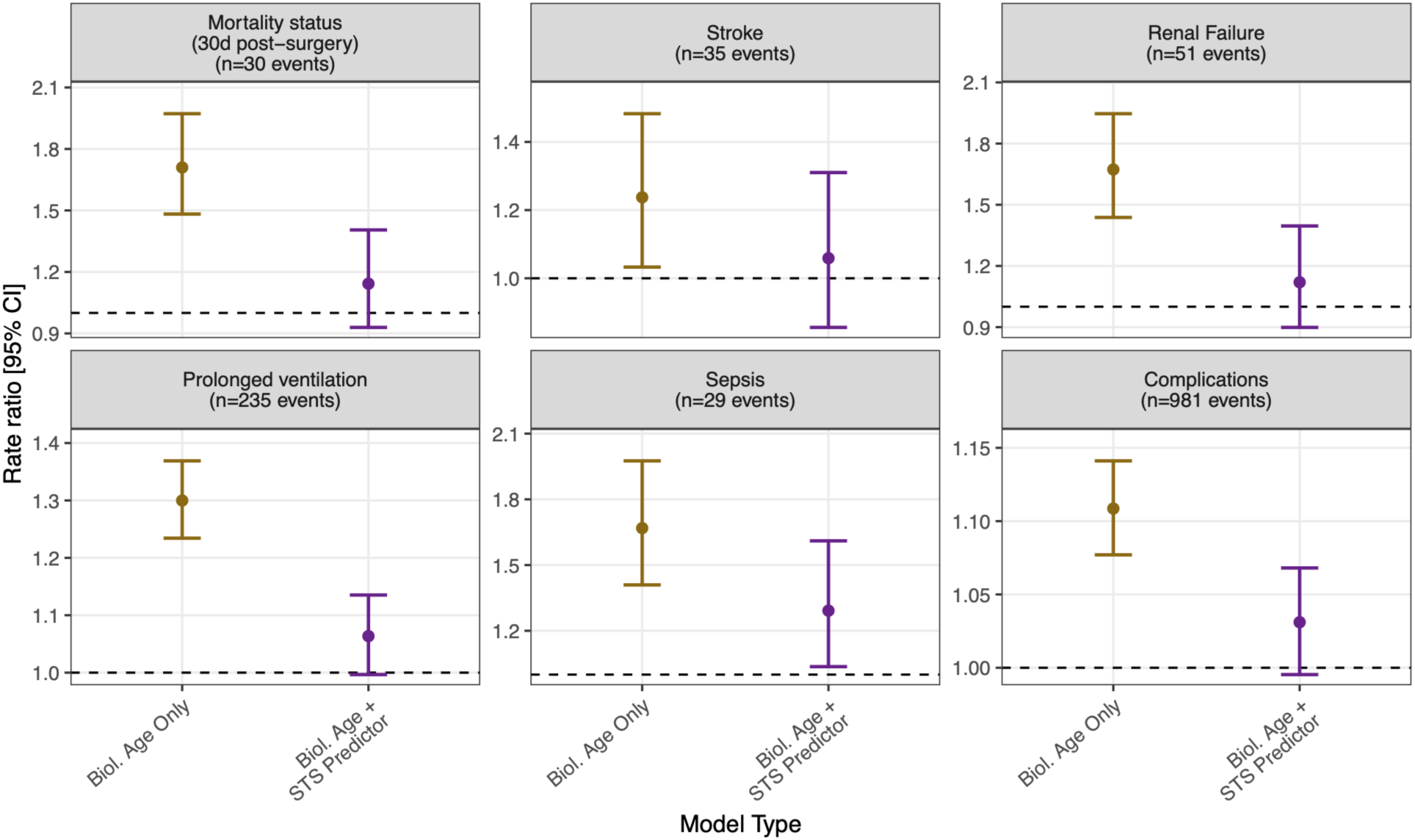
Independent predictive value of pre-operative biological age before and after adjustment for STS surgical risk scores. Models were fit separately for men (blue) and women (red) across six outcomes: 30-day mortality (n=2241), stroke (n=981), renal failure (n=932), prolonged ventilation (n=981), sepsis (n=981), and complications of surgery (n=2241). Number of events for both sexes is shown below each figure label. Each outcome was modeled twice: once with biological age alone (gold) and once with both biological age and the corresponding Society of Thoracic Surgeons (STS) predicted risk score (purple). Rate ratios (95% CIs) represent the change in risk per standard deviation increase in average 6-month pre-operative biological age, estimated from Poisson regression models adjusted for age and age² at the time of surgery. After adjustment for standardized STS predictors, pre-operative biological age only independently predicted sepsis, indicating that average 6-month pre-operative biological age does not capture additional risk information beyond established clinical models. Numbers above points denote the number of patients included in each model.

**Figure S10.**
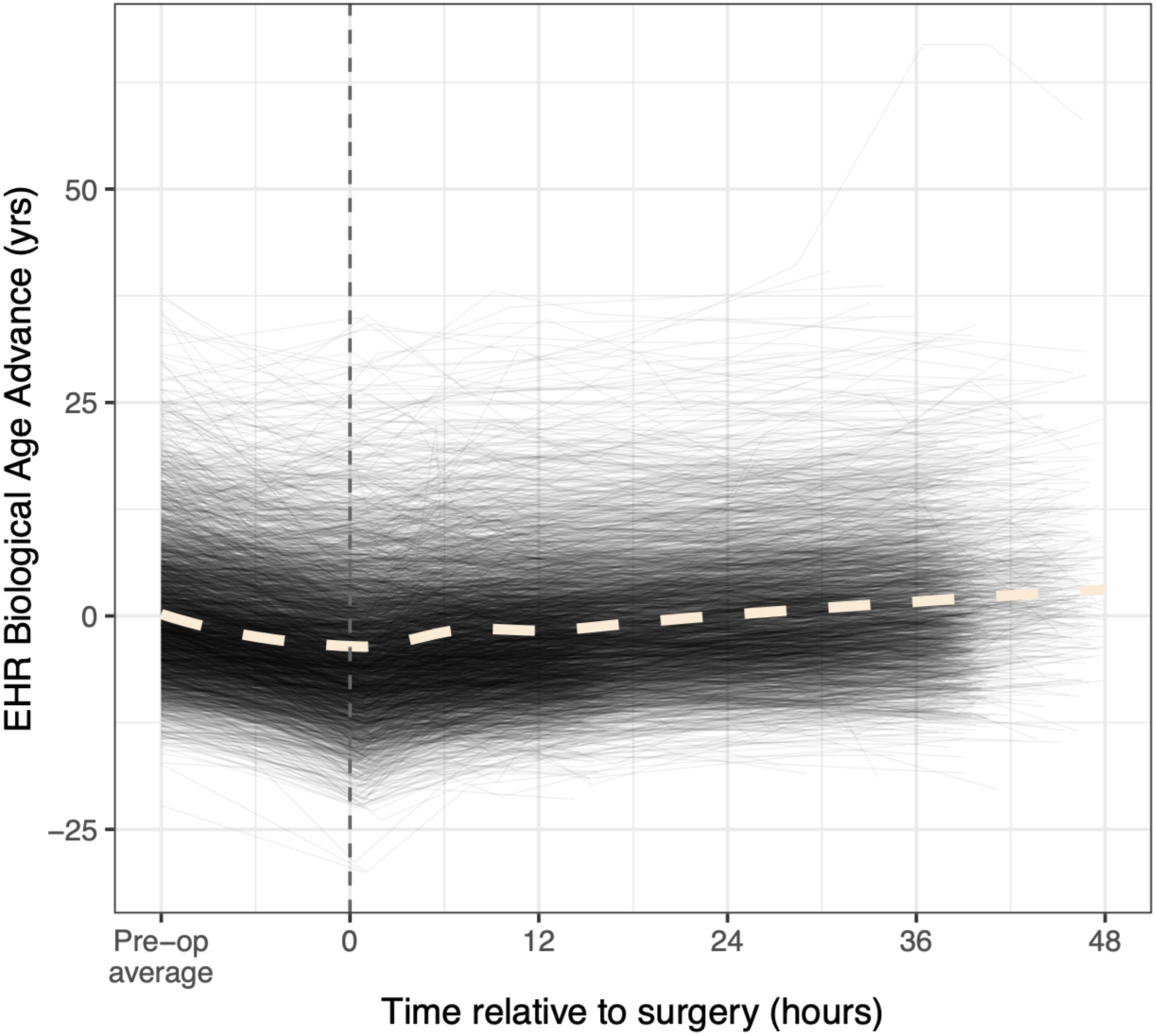
**EHR BioAge Advance immediately following surgery.** Individual trajectories show EHR BioAge Advance from the participant-specific average of all available preoperative values within six months of surgery, displayed at −12 hours as a reference, through measurements collected during the first 48 hours after surgery. EHR BioAge Advance is calculated as EHR BioAge minus chronological age, with higher values indicating more advanced biological age. The vertical dashed black line marks the time of surgery, and the dashed cream LOESS curve summarizes the overall trajectory. The preoperative average is positioned at −12 hours for visualization and does not represent the actual measurement time.

**Figure S11.**
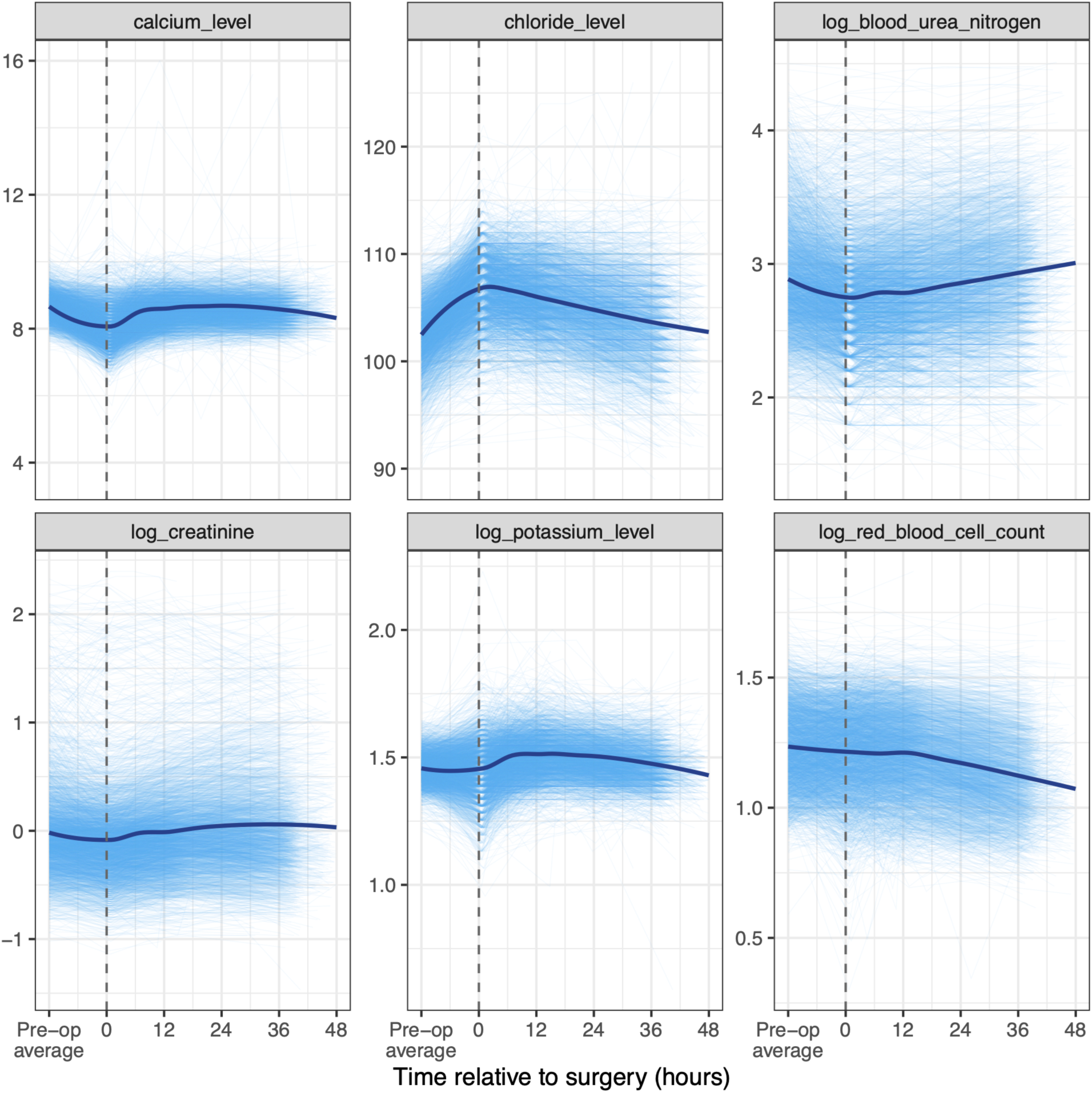
**Perioperative trajectories of calcium, chloride, blood urea nitrogen, creatinine, potassium, and red blood cell counts.** Individual trajectories show biomarker values from the participant-specific average of all available preoperative measurements values within six months of surgery, displayed at −12 hours as a reference, through measurements collected during the first 48 hours after surgery. The vertical dashed black line marks the time of surgery, and royal blue LOESS curves summarize the overall trajectory for each biomarker. Preoperative averages are positioned at −12 hours for visualization and do not represent the actual measurement times. Biomarkers are shown on separate scales. Blood urea nitrogen, creatinine, potassium, and red blood cell count are logged.

**Figure S12.**
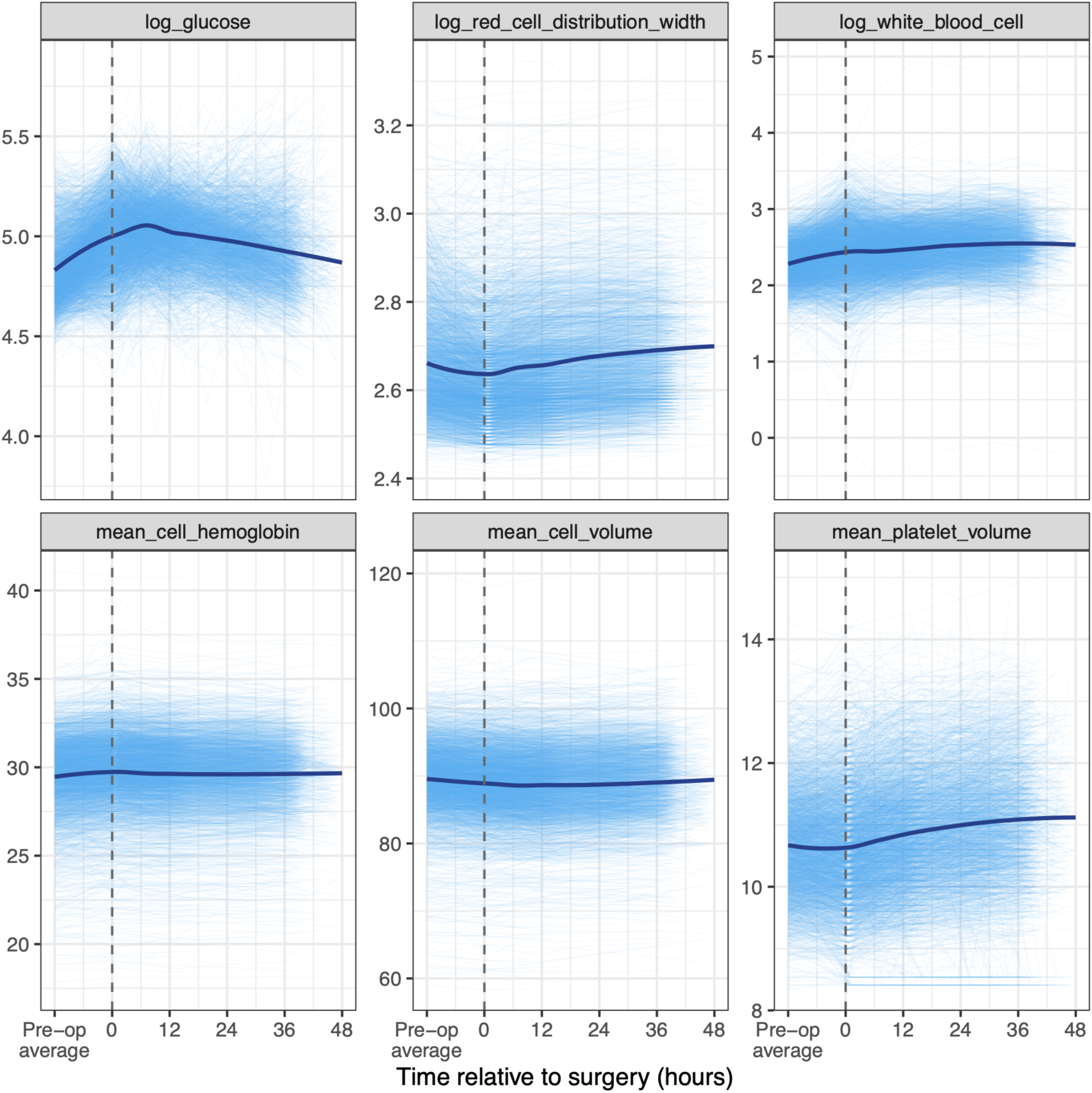
**Perioperative trajectories of glucose, red cell distribution width, white blood cell count, mean cell hemoglobin, mean cell volume, and mean platelet volume.** Individual trajectories show biomarker values from the participant-specific average of all available preoperative measurements values within six months of surgery, displayed at −12 hours as a reference, through measurements collected during the first 48 hours after surgery. The vertical dashed black line marks the time of surgery, and royal blue LOESS curves summarize the overall trajectory for each biomarker. Preoperative averages are positioned at −12 hours for visualization and do not represent the actual measurement times. Biomarkers are shown on separate scales. Glucose, red cell distribution width, and white blood cell count are logged.

**Figure S13.**
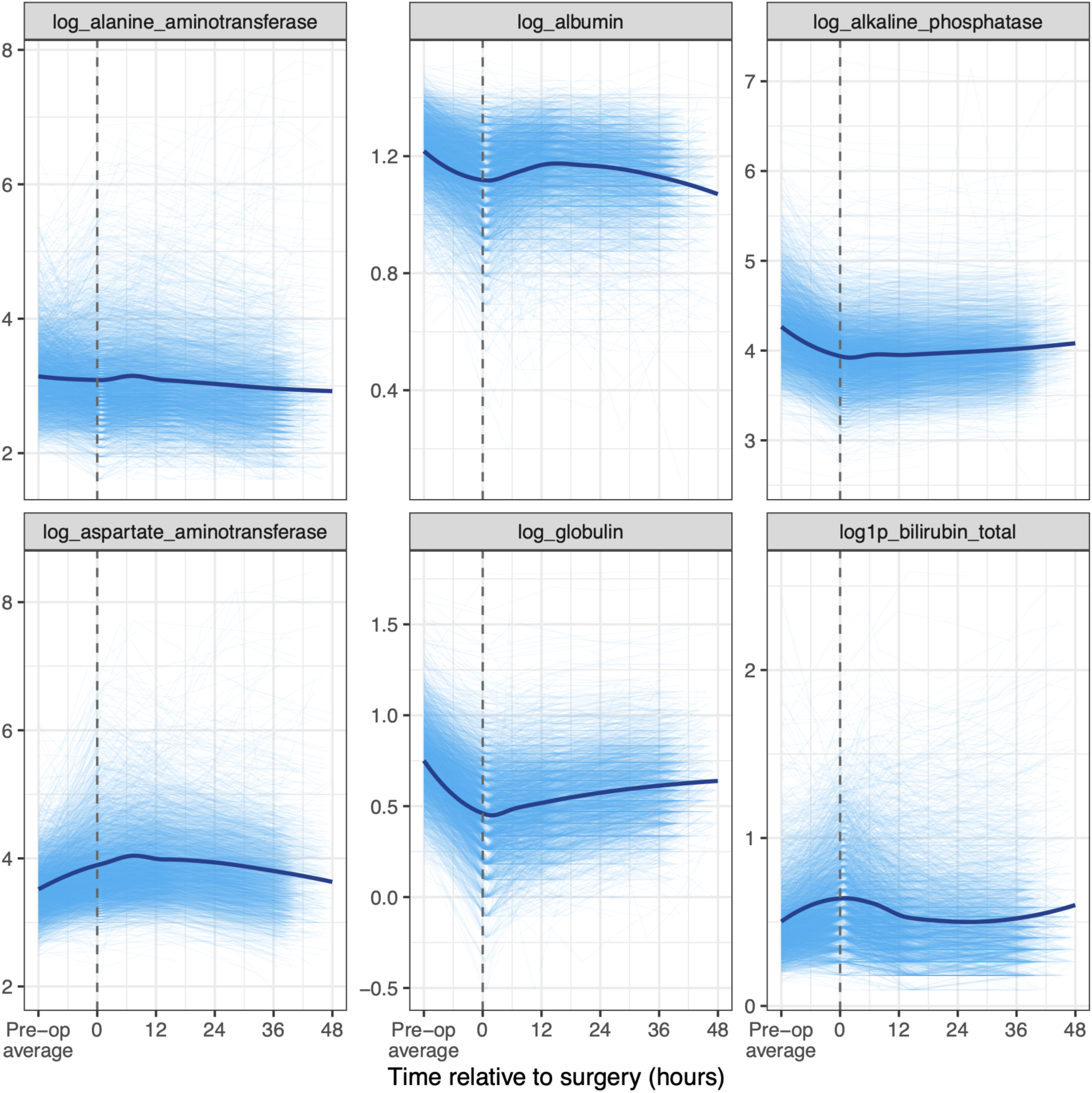
**Perioperative trajectories of alanine aminotransferase, albumin, alkaline phosphatase, aspartate aminotransferase, globulin, and bilirubin.** Individual trajectories show biomarker values from the participant-specific average of all available preoperative measurements values within six months of surgery, displayed at −12 hours as a reference, through measurements collected during the first 48 hours after surgery. The vertical dashed black line marks the time of surgery, and royal blue LOESS curves summarize the overall trajectory for each biomarker. Preoperative averages are positioned at −12 hours for visualization and do not represent the actual measurement times. Biomarkers are shown on separate scales. All measures are logged. A constant of 1 was added to bilirubin prior to logging.

**Figure S14.**
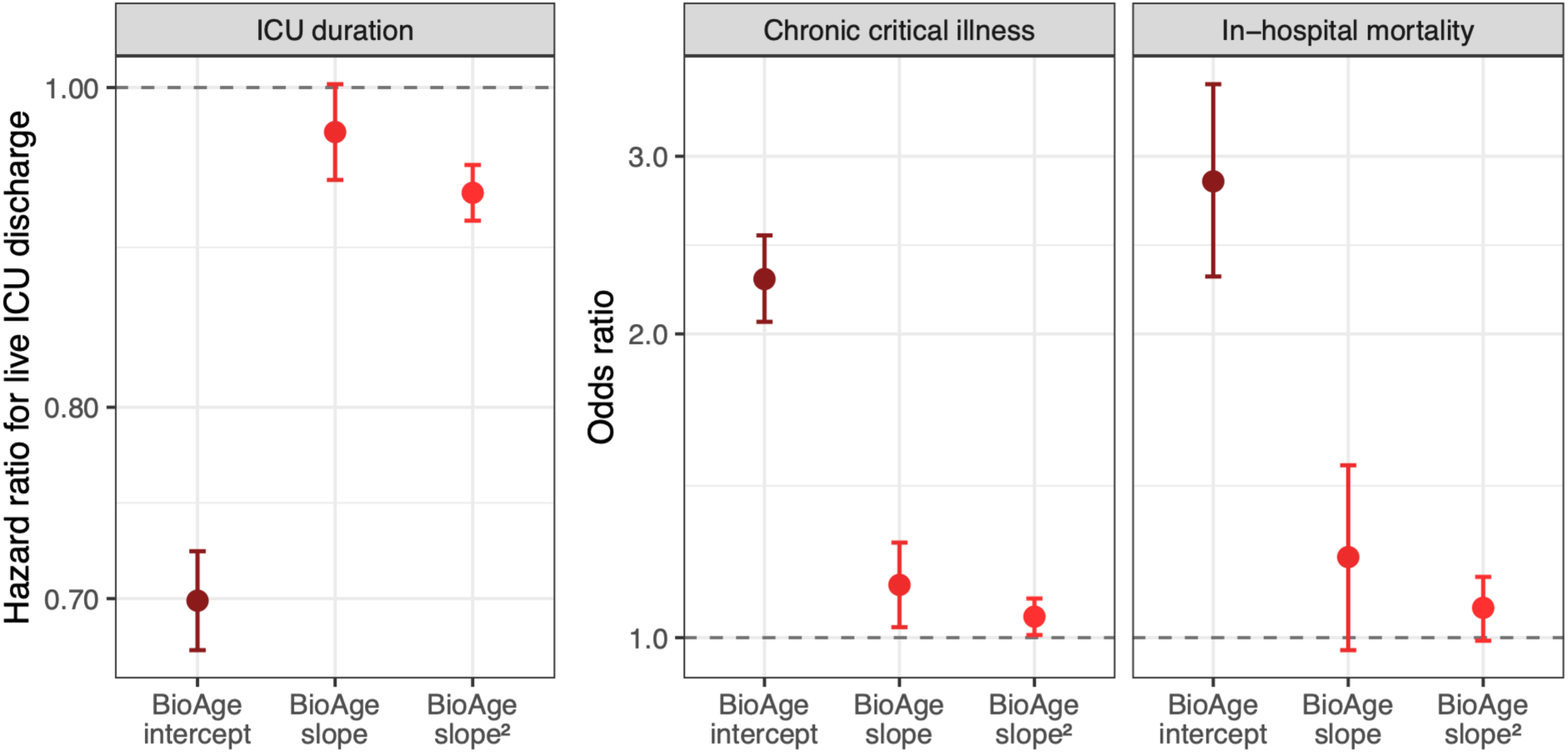
**Associations of postoperative EHR BioAge trajectories with ICU duration, chronic critical illness, and in-hospital mortality.** Participant-specific postoperative EHR BioAge intercepts and slopes were estimated from a linear mixed-effects model of repeated EHR BioAge measurements following surgery. The forest plots show mutually adjusted associations of the standardized BioAge intercept, standardized BioAge slope, and squared standardized slope with time to live ICU discharge (left), chronic critical illness, defined as ICU stay exceeding 7 days (middle), and in-hospital mortality (right). Hazard ratios for live ICU discharge were estimated using a Cox proportional hazards model; values below 1 indicate slower discharge and longer ICU stay. Odds ratios for chronic critical illness and mortality were estimated using logistic regression; values above 1 indicate greater odds of the outcome. All models included the three BioAge trajectory terms simultaneously and were adjusted for age at surgery, age squared, and sex. Points show hazard or odds ratios, error bars indicate 95% confidence intervals, and dashed lines denote the null value of 1.

**Figure S15.**
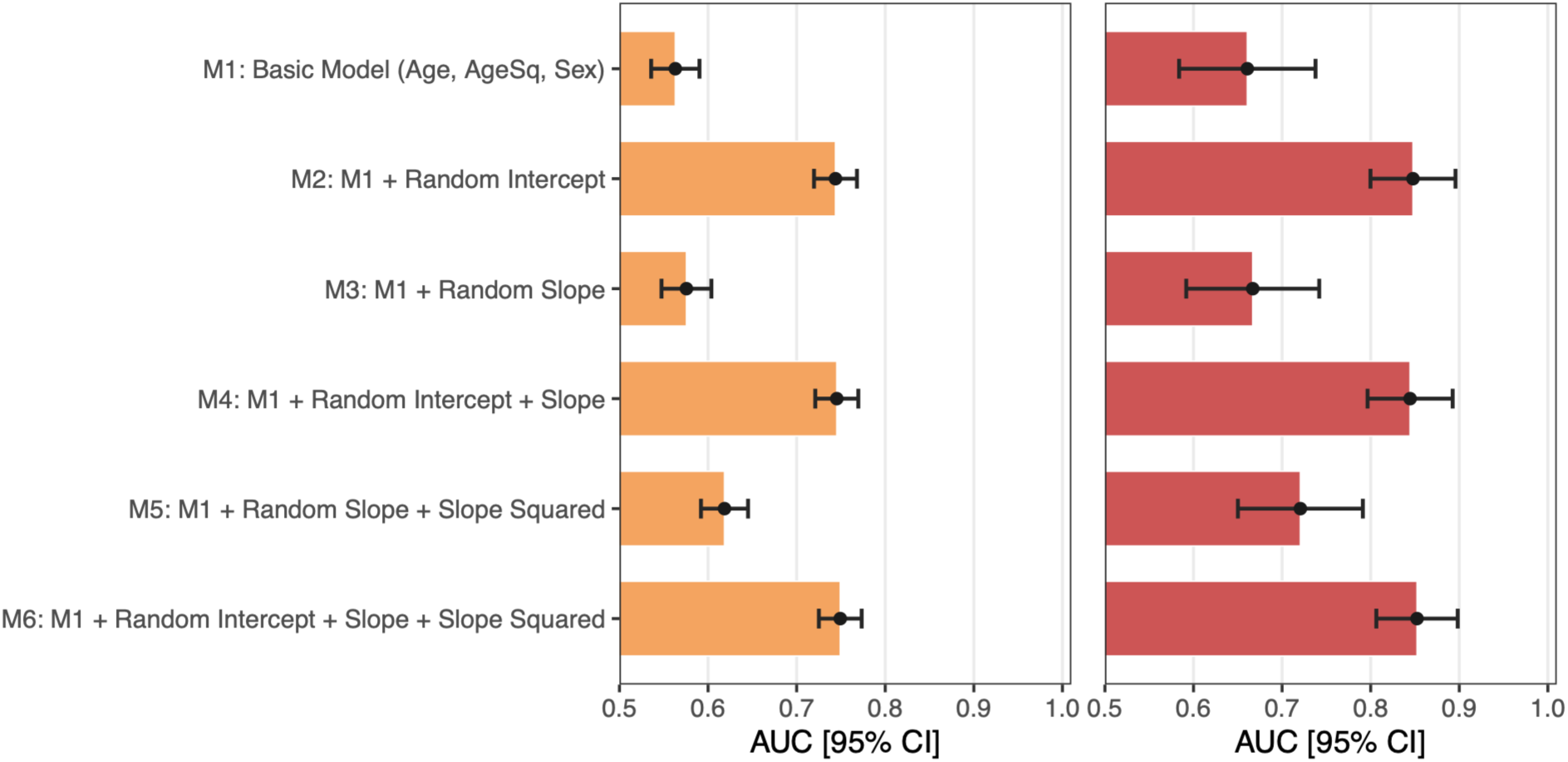
**Discrimination of postoperative EHR BioAge trajectory models for chronic critical illness and in-hospital mortality.** Areas under the receiver operating characteristic curve (AUCs) are shown for generalized linear models with a binomial distribution and logit link predicting chronic critical illness, defined as ICU stay exceeding 7 days (left, orange), and in-hospital mortality (right, red). The basic model (M1) included age at surgery, age squared, and sex. Subsequent models added the participant-specific postoperative EHR BioAge random intercept (M2), random slope (M3), random intercept and slope (M4), random slope and slope squared (M5), or random intercept, slope, and slope squared (M6). Random intercepts and slopes were estimated from a linear mixed-effects model of repeated EHR BioAge measurements obtained during the first 48 hours after surgery. EHR BioAge intercepts and slopes were standardized, and slope squared was calculated as the square of the standardized slope. AUCs were calculated from the model-predicted probabilities in the analytic sample. Black points denote AUC estimates, error bars indicate 95% confidence intervals, and colored bars extend from the chance-discrimination value of 0.50 to the estimated AUC.

