## Supplementary Methods for "Biological aging measured from electronic health records is associated with recovery following cardiothoracic surgery"

### Supplementary Methods and Results

We pulled records for all labs linked to NYP patients on February 7, 2025 and joined with Clarity component information allowing us to connect key information from each individual record to broader metadata about measures (i.e. biomarkers in panels to individual measures using LOINC codes).[1] We searched all biomarkers belonging to the three most common panels in our dataset: a basic metabolic panel, a hepatic function panel, and a CBC platelet without differential panel. We then used associated LOINC codes to re-search for any of the 22 individual biomarkers that were measured outside of the standard panel. For each biomarker, we checked for skewness using Fisher-Pearson standardized third moment and biomarkers with skewness >0.5 were logged. We added 1 to bilirubin levels prior to logging due to many small values.

We assembled and harmonized laboratory and clinical data from the NYP EHR to evaluate associations between biological age and short-term surgical outcomes. Complete demographic, operative, post-operative, and discharge information were available for n=4,313 (**Fig. 1**). Key outcome variables and covariates included in this analysis, their names and definitions as defined in the STS Adult Cardiac Surgery Database (ACSD) Training Manual are provided in **Table S1**.

From a total of 9,693,200 lab records, we searched analytes belonging to a basic metabolic panel, a hepatic function panel, and a CBC platelet without differential panel, resulting in 2,088,538 individual labs across 22 analytes. After our second pass search using associated LOINC codes our dataset consisted of 2,631,573 labs across all n=4,313 patients. Number of individual records and patients for each lab can be found in **Table S2**. In total, there were 71,947 instances where all 22 labs were available at the same time for the same person (15,157 instances among n=2781 patients prior to surgery; 56,790 instances among n=4297 patients after surgery). **Figure 1** shows the number of labs and patients through processing. Analytes with skewness >0.5 were logged. Analytes and their transformations are provided in **Table S3**.

##### *National Health and Nutrition Examination Survey (NHANES).*

We used NHANES III to train an adapted biological age model restricted to biomarkers available in the NYP dataset. NHANES is an ongoing nationally representative, cross-sectional survey conducted by the US Centers for Disease Control and Prevention. NHANES administers questionnaires during in-home interviews and conducts health examinations, including blood draws, in a mobile examination center. Details of recruitment procedures and study design are available from the Center for Disease Control and Prevention (https://www.cdc.gov/nchs/nhanes/index.htm). We compiled demographic, socioeconomic, and functional performance; biomarker; and mortality data from adults aged 20–90 years participating in the NHANES III (1988–1994).

##### *Biomarker processing, harmonization, and biological age construction in NHANES.*

Using the list of biomarkers and skewness information from our EHR dataset, we created a new dataset using laboratory, exam, and mortality data in NHANES III. Corresponding biomarkers in NHANES were identified, transformed, and scaled (mean=0, standard deviation =1), and observations >5 standard deviations from the sex-specific mean were removed. Details on biomarker measurements are available from the NHANES website (https://www.cdc.gov/nchs/nhanes/index.htm).

We computed blood-chemistry measures using the BioAge R package[2]. The biological age algorithm is derived from multivariate analysis of mortality hazards.[3,4] The original biological age algorithm was constructed from elastic-net Gompertz regression of mortality on 42 biomarkers in the NHANES III.[5] To optimize our list of biomarkers and maximize our sample size, we first fit an elastic net Cox regression predicting mortality (n=11,969 with 4,753 deaths) with all 22 scaled biomarkers, age, and sex, using 10-fold cross-validation with alpha set to 0.5.[6] The best elastic net Cox regression fit in NHANES included 18 of the original 22 analytes. The analytes selected in this model and their coefficients are provided in **Table S4**.

We then applied the *BioAge_calc* function from the BioAge package to individuals with complete data for chronological age and for all 18 biomarkers identified in the prior elastic net Cox regression. In this step, we fit sex-specific Gompertz proportional hazards model relating mortality risk to chronological age and the unscaled biomarkers, allowing us to construct a new measure of biological age tailored to biomarkers available in our EHR data (biological age). In sample, biological age was highly-correlated with age in both men (r2 men=0.93; r2 women=0.96; **Figure S1**) and was strongly predictive of mortality in the training data ($p<0.0001$ for both sexes). Sex-specific model parameters are shown in **Table S5**.

##### *Biological age measure validation*

In the third step, we validated our algorithm by comparing its prediction of morbidity and mortality with Levine's original PhenoAge algorithm in data from NHANES IV (1999-2018). We derived NHANES IV analytes used for calculating EHR biological age by searching all datasets between 1999-2025 for EHR laboratory analytes using any partial word matches and manually curating to include only relevant assays, sample tissue type, and units. We then pulled datasets containing focal analytes directly from the NHANES website using the *nhanesa* R package[7]. We harmonized analyte names across waves, averaged any duplicated measures within waves, and combined all years into one dataset. We derived sex and age at screening from a separate data pull of demographic datasets. We logged all analytes, stratified by sex, and scaled in standard deviations from the mean. Values >5 standard deviations from sex-specific mean were removed. We next transformed analytes using the same methods used for NHANES III and in accordance with the distributions of these measures in the EHR. We then split the data by sex and applied the sex-specific EHR biological age algorithm derived in NHANES III to each sex separately.

For survival analyses, EHR biological age in NHANES IV was joined with updated mortality data from 2019. For direct comparisons with PhenoAge we then joined these data with previously curated NHANES IV dataset from Kwon and Belsky[2], for a total of n=33,251 participants in survival analysis.

EHR biological age in NHANES IV showed strong correlation with chronological age (r=0.97; **Figure S2**), and a strong correlation with PhenoAge (r=0.72; **Figure S3**). Mortality prediction of EHR biological age was similar to PhenoAge (HR: 1.45, 95%: 1.41-1.48 for EHR Biol. Age vs. HR: 1.49, 95%CI: 1.46-1.52 for PhenoAge), including when stratified by sex, race, and age (**Table S6**).

##### *Biological age projection in NYP EHR.*

We projected NHANES-derived biological age algorithm into EHR data using the biomarkers and coefficients selected in NHANES. We grouped the 18 biomarkers selected in the NHANES biological age measure generation step by patient per timepoint in the EHR dataset. If patients had numerous tests for same biomarker run at the same time, we took the average of all tests. If patients were missing values for 4 or fewer biomarkers, we carried out an imputation step where we substituted missing biomarker values with the sex-specific biomarker mean from NHANES. We then implemented the biological age algorithm on each sex separately and scaled each measure to have a sex-specific Mean=0 and $SD=SD_{NHANES}$, the equivalent to one standard deviation for the corresponding sex in the NHANES training data.

#### Part I: Pre-operative biological age and post-operative outcomes

##### Pre-operative biological age time windows.

Available pre-operative laboratory records spanned a wide temporal range, with measurements dating back up to 13.75 years prior to surgery (mean = 337.7 days; SD = 651), although most samples were collected shortly before surgery (median = 9.8 days).

To define a pre-operative biological age exposure that was both biologically meaningful and broadly available across patients, we evaluated the stability of biological age estimates across alternative pre-operative time horizons. Specifically, we constructed four candidate biological age summaries for each patient: (i) the average of all available pre-operative measurements, (ii) the average of measurements obtained within 1 year of surgery, (iii) the average of measurements obtained within 6 months of surgery, and (iv) the average of measurements obtained within 3 months of surgery. Within each window, biological age values were averaged across all qualifying timepoints.

We compared these candidate windows with respect to (i) the number of biological age observations per individual, (ii) correlations between biological age estimates across windows, (iii) correlations with chronological age, and (iv) the number of patients and outcome events available for downstream analyses. To ensure that window choice did not materially influence inference, we additionally verified that associations between biological age and postoperative outcomes were consistent across windows when evaluated using identical model specifications (described below). Based on these considerations, we selected the average biological age calculated from measurements obtained within 6 months prior to surgery as the primary exposure for all subsequent pre-operative analyses. This window balanced proximity to surgery, biological relevance, and data completeness.

##### *Distribution and timing of pre-operative biological age*

We first focused our analysis on biological age measurements calculated prior to surgery. Laboratory records used to compile biological age dated back to as early as 13.75y prior to the date of surgery (mean=337.7 days, sd=651), but most samples were taken within 10 days of surgery (median=9.8 days). We averaged biological age from all pre-operative timepoints, those taken within a year, within 6 months of surgery, and within 3 months of surgery and compared them. Median number of biological age measurements for all pre-operative windows were 2 times, except the 3-month pre-operative window where the median number of biological age measurements was once (**Fig. S4**). All time windows were highly correlated with each other ($r2>0.92$, **Figure S5**), and were correlated with chronological age (pearson $r > 0.8$; **Table S7**), and had a comparable number of patients and events in outcomes of interest (**Figure S6**). To balance biological signal with data completeness, we focused our analyses on the average of all measurements taken within 6 months of surgery. Sex-specific histograms and scatterplots of average 6-month biological age with chronological age in our EHR data are shown in **Figure 2**.

##### *Assessing the predictive contribution of pre-operative biological age*

We evaluated associations between pre-operative biological age and short-term postoperative outcomes using generalized linear models estimating risk ratios. Models were fit using a Poisson family with a log link and included age at surgery, age squared, sex, and either biological age (scaled to a sex-specific mean of 0 and standard deviation equal to the sex-specific standard deviation in the NHANES training data) or biological age in combination with the Society of Thoracic Surgeons (STS) risk score corresponding to the outcome of interest (mortality, stroke, renal failure, prolonged ventilation, sepsis, or composite complications).

##### Society of Thoracic Surgeons (STS) Risk scores.

Risk score predictions for mortality, stroke, renal failure, prolonged ventilation, sepsis, and composite complications were available for a subset of patients. These estimates are generated by the Society of Thoracic Surgeons (STS) risk models, which integrate a detailed set of pre-operative clinical characteristics, comorbidities, physiologic measures, and procedural factors (e.g., age, cardiac function, disease severity, hemodynamic instability, and type/urgency of surgery) to estimate patient-specific operative risk. The development, validation, and clinical rationale for these models have been described in detail elsewhere.[8,9] A screenshot of the online risk calculator interface is shown in **Figure S7**. Key risk scores and their names and definitions, as described in the STS Adult Cardiac Surgery Database (ACSD) Training Manual are provided in **Table S1**.

##### *Comparative Discrimination of pre-operative biological age and STS Risk Scores*

To compare the discriminative performance of biological age with established surgical risk predictors, we fit logistic regression models predicting each of the six short-term postoperative outcomes. For each outcome, we calculated the area under the receiver operating characteristic curve (AUC) in (i) the full sample of patients with biological age and outcome data and (ii) the subset of patients with both biological age and the corresponding STS predicted risk score available (‘overlap sample’). Models included: (1) average 6-month pre-operative biological age alone, (2) the log-transformed STS risk score alone, and (3) a combined model including log(STS score), biological age, age, age$^2$, and sex. AUCs and 95% confidence intervals were estimated using the pROC package in R.[10]

#### Part II: Post-operative biological age and post-surgical recovery

##### *Post-operative cohort definition*

We examined whether baseline post-operative biological age and short-term post-operative trajectories in biological age during the first 48 hours after surgery were associated with post-surgical recovery. Outcomes included (i) total time spent in the intensive care unit (ICU), (ii) prolonged ICU stay (>7 days), reflecting transition from acute to chronic critical illness, and (iii) in-hospital mortality. To ensure interpretable trajectory estimation, analyses were restricted to patients with a single continuous ICU stay; patients stepped down from ICU and later readmitted (n = 132) were excluded. Hospital death was restricted to patients who were discharged alive and their last known status was alive, or those who died in the hospital. Patients who were discharged to hospice or who died after discharge were excluded. ICU length of stay was taken from the time of exit from the operating room to the time of discharge from the ICU. Key variables included in this analysis, their names and definitions as defined in the STS Adult Cardiac Surgery Database (ACSD) Training Manual are provided in **Table S1**.

Because biological age may convey prognostic information both through its overall post-operative level and through its short-term dynamics, we characterized early post-operative biological age as an average biological age across the first 48 hours after surgery, and as a patient-specific trajectory capturing the rate and shape of biological age change during this period.

##### *Estimation of average and trajectory-based post-operative biological age*

###### Average post-operative biological age

To characterize each patient's overall biological age burden during the immediate post-operative period, we computed an average post-operative biological age using all available biological age measurements obtained within the first 48 hours following surgery. This summary measure parallels the pre-operative biological age exposure and provides a simple, time-agnostic characterization of early post-operative biological aging.

###### Post-operative biological age trajectories

To characterize patient-specific trajectories of biological age acceleration during the first 48 hours following surgery, we fit a linear mixed-effects model.[11] Biological age acceleration was modeled as a function of time since surgery (in days), age at surgery (linear and quadratic terms), and sex, with a random intercept and random slope for time at the patient level. The random intercept represents each patient's baseline deviation in post-operative biological age acceleration, while the random slope represents the patient-specific rate of change over time. The model can be written as:

$$\text{Biological age advance}_{it} = \beta_0 + \beta_1\,\text{TimeDays}_{it} + \beta_2\,\text{AgeAtOR}_i + \beta_3\,\text{AgeAtOR}_i^2 + \beta_4\,\text{Sex}_i + u_{0i} + u_{1i}\,\text{TimeDays}_{it} + \varepsilon_{it}$$

where $u_{0i}$ and $u_{1i}$ represent the patient-specific random intercept and random slope for time, respectively, and $\varepsilon_{it}$ is the residual error term.

From this model, we extracted patient-level random intercepts and random slopes for time. These quantities were standardized (mean = 0, SD = 1) and used as predictors in downstream outcome models. In models allowing non-linearity in trajectories, we additionally included the squared standardized slope term to capture curvature in biological age change during the early post-operative period

##### *Post-operative outcomes and statistical models*

###### ICU length of stay (time to event analysis)

To evaluate associations between biological age dynamics and total ICU length of stay, we fit five Cox proportional hazards models predicting time to ICU discharge (in hours).[12] All models adjusted for age at surgery (linear and quadratic terms) and sex, and differed only in their specification of biological age predictors: i) random intercept only (baseline biological age); ii) random slope only (rate of biological age change); iii) random slope and intercept only; iv) random slope and squared-slope, and; v) random intercept, random slope, and squared-slope. Hazard ratios (HRs) with 95% confidence intervals (CIs) are reported.

###### Prolonged ICU stay (>7 days)

To assess whether baseline biological age and early biological age trajectories predicted progression from acute to chronic critical illness, we fit five logistic regression models with ICU stay longer than 7 days as the binary outcome. Covariate adjustment and biological age model specifications mirrored those used in the Cox models described above. Results are reported as odds ratios (ORs) with 95% CIs.

##### In hospital mortality

To evaluate associations with mortality during the index hospitalization, we fit five logistic regression models with in-hospital death as the outcome. Models included age at surgery (linear and quadratic terms), sex, and the same biological age specifications used for ICU outcomes (models 1–5). Odds ratios with 95% CIs are reported.

###### *Predictive performance*

For the two binary outcomes (prolonged ICU stay and in-hospital mortality), we evaluated predictive performance by generating predicted probabilities from each biological age model and from a base model including only age (linear and quadratic terms) and sex. Model discrimination was compared using the area under the receiver operating characteristic curve (AUC), estimated using the pROC[10] package in R.

###### *Distribution and timing of post-operative biological age*

The average post-operative ICU stay was 10.0 days (sd=17.4 days). ICU stay ranged from 1.4 hours to 183 days. A total of n=3991 patients had biological age measures within the first 48 hours and post-operative ICU discharge information. Of these, n=494 (12.3%) patients spent over 7 days in the ICU, meeting the unit definition for chronic illness. From n=3968 patients with unambiguous survival data at last follow-up (i.e. patient either died in hospital or were discharged alive and remained alive at the last follow-up), n=56 died in hospital.

#### Supplementary Methods References

12. Therneau, T. A package for survival analysis in R. (2020).
